# The Generative AI Meta-Evaluation (GAME) Study Framework: Global, Regional, and Country-Specific Unequal Difficulty of High BMI Intervention

**DOI:** 10.64898/2026.03.23.26349046

**Authors:** Chen Sun, Chuanwu Liu, Wenhua Lv, Wei She, Siyu Wei, Haiyan Chen, Junxian Tao, Jing Xu, Tianyang Lei, Qiong Wu, Yuan Xu, Ning Wang, Yan Guo, Qinduo Ren, Chang Wang, Songlin Lu, Zhenwei Shang, Chi Yan, Jingyang Hu, Tianyu Zhou, Qinghua Liu, GRD 2025 global high BMI and intervention Consortium, Mingming Zhang, Hongchao Lyu, Yongshuai Jiang

## Abstract

**Background:** High body mass index (BMI) presents a serious and ongoing global health challenge. However, the difficulty of high BMI intervention has not yet been systematically evaluated.

**Methods:** We developed a Generative Artificial Intelligence Meta-Evaluation (GAME) framework, which integrated 18 indicators from 4 dimensions, including “Macro-System Level”, “Socio-Cultural Level”, “Community-Family Level”, and “Individual Level” to evaluate the difficulty of high BMI intervention across 226 locations. The GAME framework applies 8 leading AI models to generate intervention difficulty scores (IDS) of each indicator on a scale from 1 to 5, with higher scores indicating greater difficulty. Meta-analysis was conducted to derive combined scores, evaluate the heterogeneity and sensitivity. Final intervention difficulty scores were calculated as the weighted sum of all 18 indicators. Additionally, SHapley Additive exPlanation (SHAP) values were used to evaluate the importance of each indicator in determining the intervention difficulty.

**Results:** The global difficulty of high BMI intervention shows significant imbalance. Norway (IDS = 1.48) exhibited the easiest intervention, while Yemen (IDS = 4.56) faced the greatest challenge. Regions such as Western Europe, Australasia, and High-income Asia Pacific showed lower intervention difficulty, reflecting there are mature public health frameworks, supportive social-cultural environments for healthy lifestyles, and high levels of health awareness. On the contrary, countries in North Africa and Middle East, South Asia, Oceania, and Sub-Saharan Africa faced higher intervention challenges, suggesting the need for long-term, collaborative efforts from multiple sectors. Among the 18 indicators, “Cognition and Awareness” has the most significant impact on intervention difficulty, with the SHAP value of 31.03, followed by “Family life and cognitive patterns” (18.08) and “Health Care System” (11.7). Furthermore, the IDS for high BMI was significantly correlated with Socio-Demographic Index (SDI). Higher SDI values were associated with easier interventions. Finally, the independent external empirical verification demonstrated high consistency between intervention difficulty and increase in annual prevalence of obesity, population mean BMI, and national policies. It supported the GAME framework to characterize global heterogeneity in high BMI intervention challenge. Global results were freely available at http://www.deepburden.com/high-bmi.

**Conclusion:** The difficulty of high BMI intervention varies widely across countries and regions, highlighting the need for comprehensive strategies and governance to address the growing health issue effectively.

## Introduction

Overweight and obesity is a major health crisis, presenting a significant and ongoing threat to global health progress [1–5]. In 2021, these conditions contributed to 3.71 million deaths and 129 million disability-adjusted life-years (DALYs) [6]. In the past two decades, the global age-standardised DALY rates attributable to overweight and obesity increased by more than 15% [6]. High body mass index (BMI) has been one of the top risk factors with the steepest increase in attributable burden [7–10].

A major challenge is implementing intervention policies to curb the rising rates of overweight and obesity [11–13]. However, the World Health Organization reports that in 2021, 40% of nations had implemented an operational policy, strategy, or action plan to address overweight and obesity. But the policy coverage dropped to below 10% in low-income countries [6]. For examples, the UK and the USA implemented the taxation of sugar-sweetened beverages and New York City’s ban on artificial trans fats [6, 14, 15]. On the contrary, evidence for effective obesity intervention implementation remains scarce in low– and middle-income countries [6, 16]. The difficulty of high BMI intervention shows an inequality on a global scale.

Recently, generative Artificial Intelligence (AI) has made remarkable progress [17–21]. GPT, a generative transformer model, was developed based on the extensive and multiple dataset comprising diverse online texts, such as articles, books, and websites across various subjects and languages [22–24]. DeepSeek supports continuous learning by leveraging publicly available open-source datasets, improving its adaptability to rapidly evolving knowledge and scientific reasoning [25–27]. Such AI systems hold promise for various healthcare applications, including research acceleration, clinical decision support, medical education, and so on [28–30]. This raises the question of whether AI models could be employed to integrate data from various sources, thereby accurately assessing the difficulty of high BMI intervention.

In this study, we applied 8 large AI models to evaluate the difficulty of high BMI intervention in 226 locations. We developed a Generative AI Meta-Evaluation (GAME) framework (Fig. 1) to calculate the intervention difficulty scores according to 18 key indicators from 4 dimensions: “Macro-System Level”, “Socio-Cultural Level”, “Community-Family Level”, and “Individual Level”. The influence of Socio-Demographic Index (SDI) on the difficulty of high BMI intervention was evaluated. Furthermore, we verified the reliability of results based on both independent external empirical baselines, including increase in annual age-standardised prevalence of obesity, population mean BMI, and national policies, and internal heterogeneity and sensitivity analysis.

**Figure 1.**
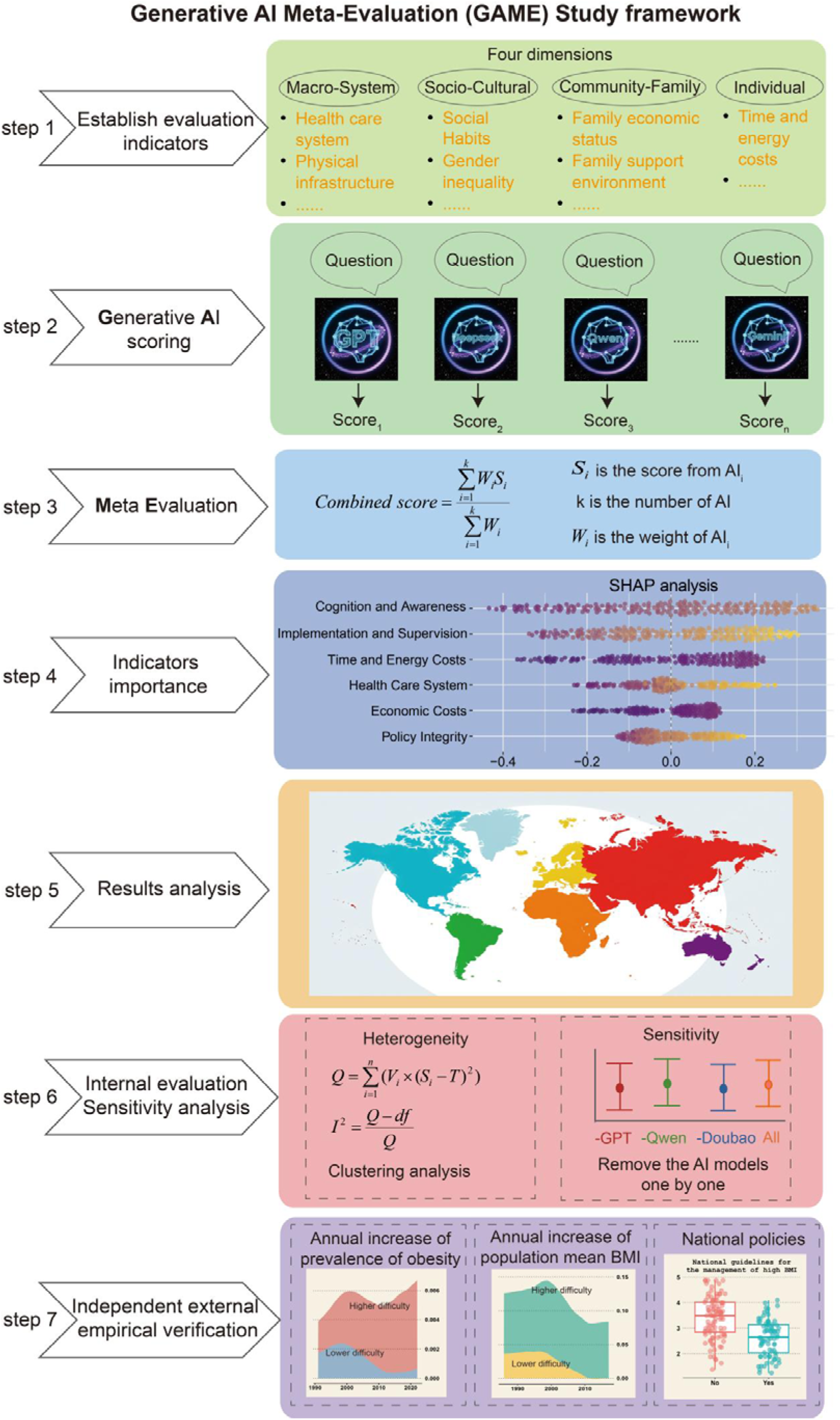
Overview of the GAME framework.

## Results

### Global, regional, and national unequal difficulty of high BMI intervention

The results revealed significant regional and national differences in the difficulty of intervening in high BMI. At the country level, the mean intervention score of high BMI was 3.15, with Norway achieving the lowest score of 1.48, and Yemen representing the highest at 4.56. Regions such as Western Europe, Australasia, High-income Asia Pacific and High-income North America exhibited lower intervention difficulty, while countries in North Africa and Middle East, South Asia, Oceania, and Sub-Saharan Africa faced higher intervention challenges (Fig. 2, Supplementary Table 1).

**Figure 2.**
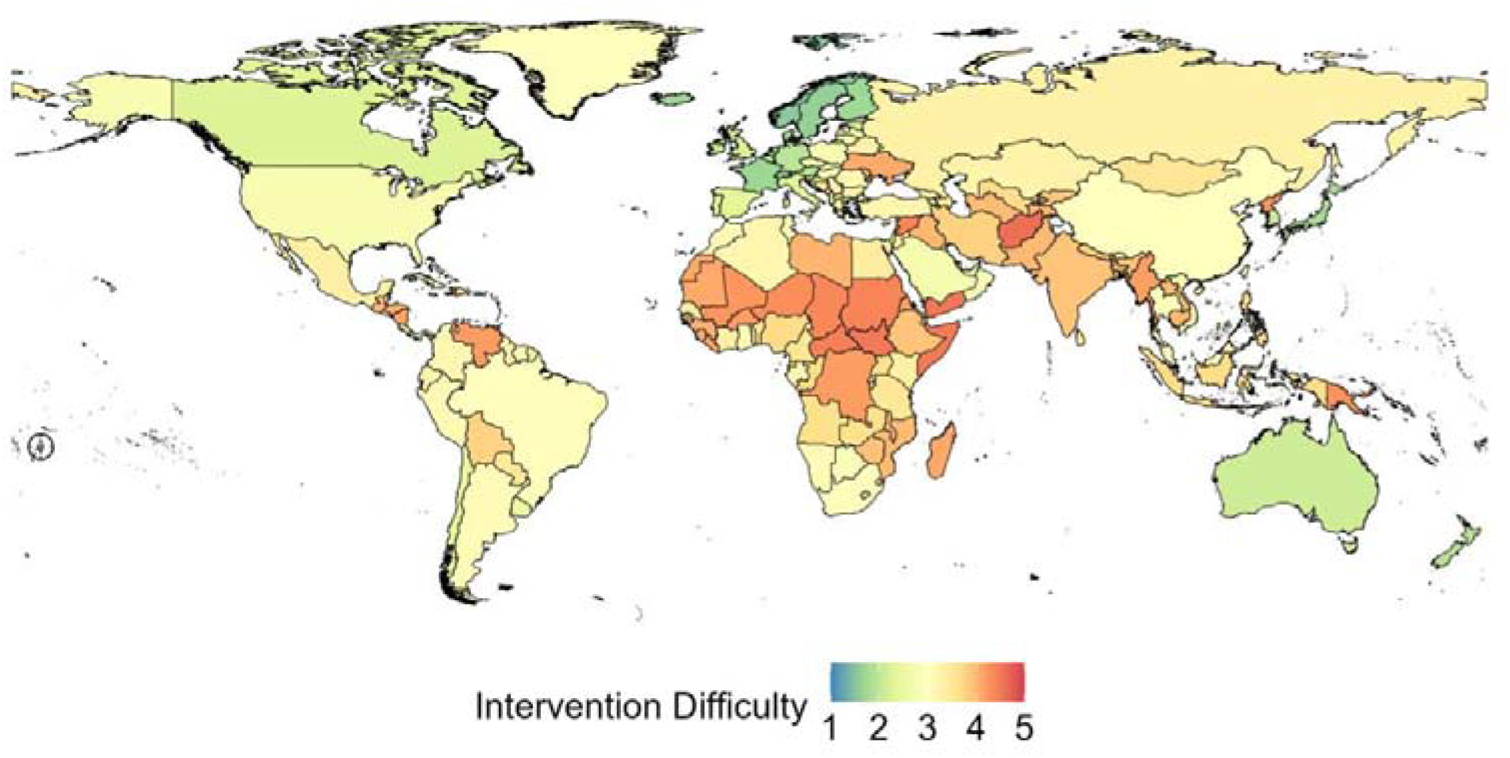
Global difficulty of high BMI intervention.

In Western Europe, the mean intervention score of high BMI was 1.96, with countries like Norway (1.48), Sweden (1.51), Iceland (1.56), and Denmark (1.64) showing the lowest difficulty. These countries benefit from strong national public health frameworks, a social culture that supports healthy lifestyle, and the medical system focused on prevention. For example, Norway has implemented high sugar taxes, mandatory nutrition labels, and universal prevention-oriented primary care. In addition, there is a strong population awareness of obesity risks (Fig. 3B). Australasia, which includes Australia (1.95) and New Zealand (2.01) followed closely, with developed economies and rich health resources, though processed foods, such as dairy products and processed meats, are common in the diet (Fig. 3D). High-income North America had a mean intervention score of 2.55, including Canada (2.08) and United States of America (3.02). Despite high income and high health awareness, the United States of America faces challenges due to an uneven distribution of health resources, strong lobbying by unhealthy industries, contributing to widespread addiction to high sugar and high fat foods (Fig. 3C).

**Figure 3.**
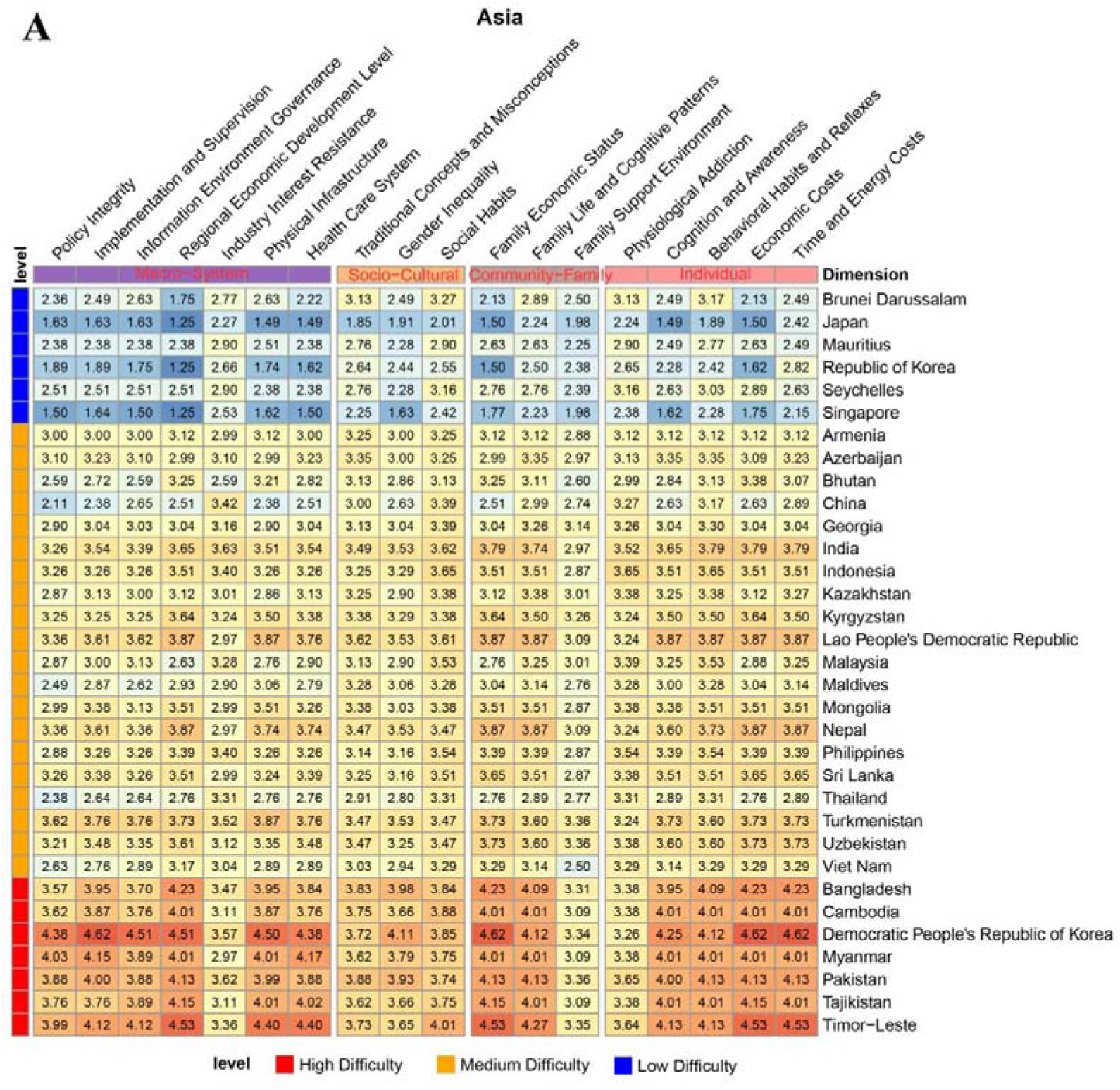
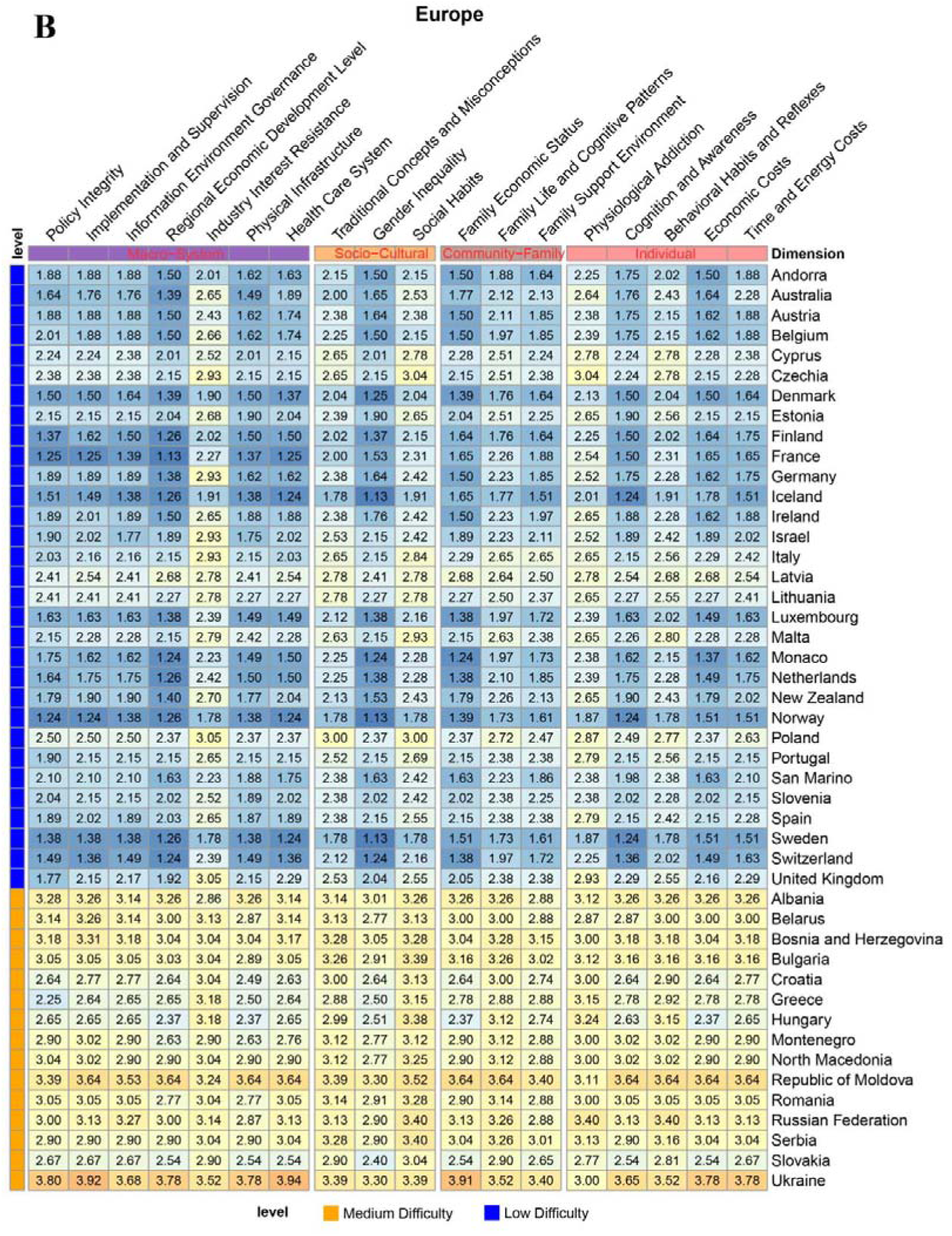
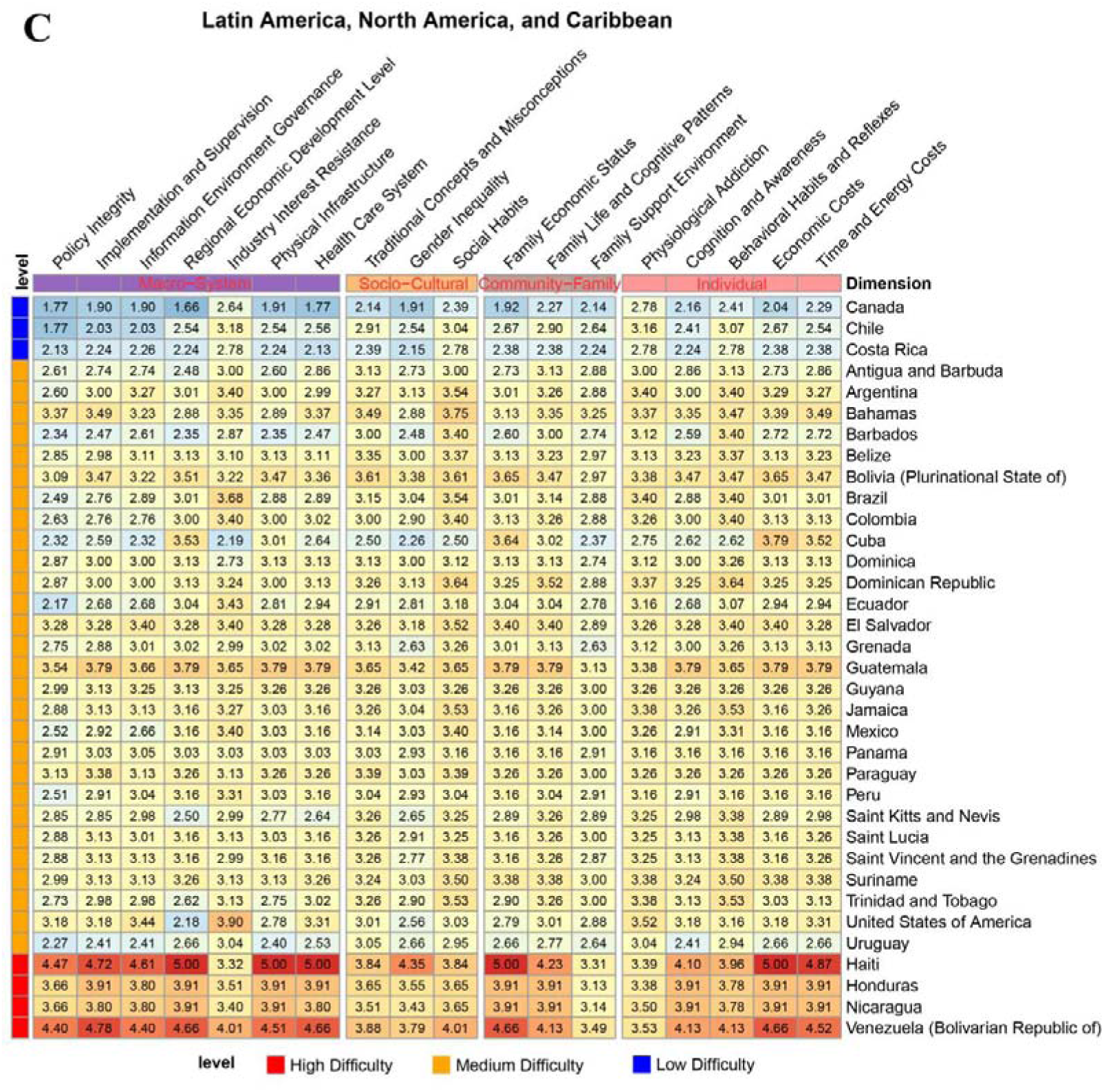
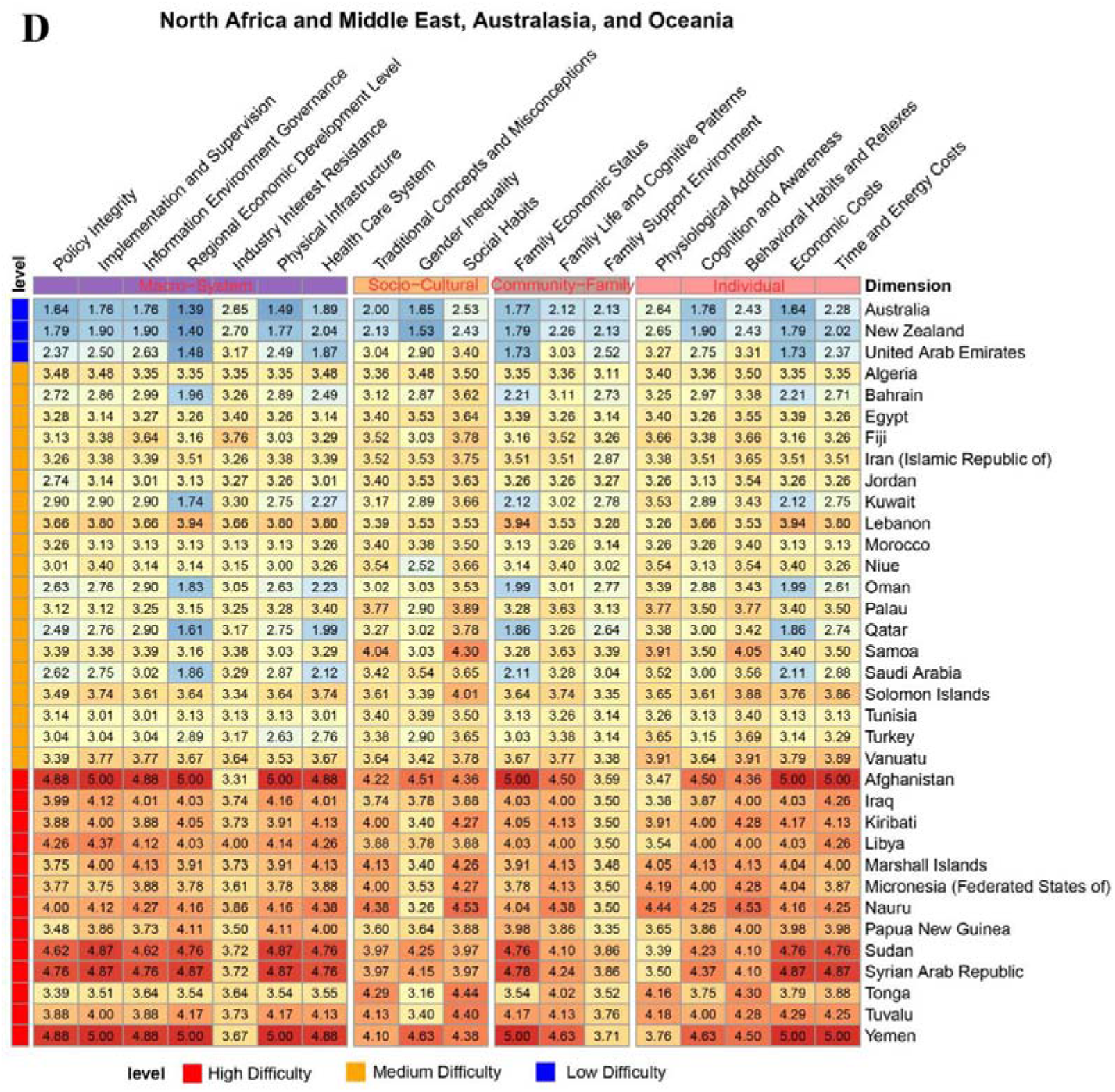
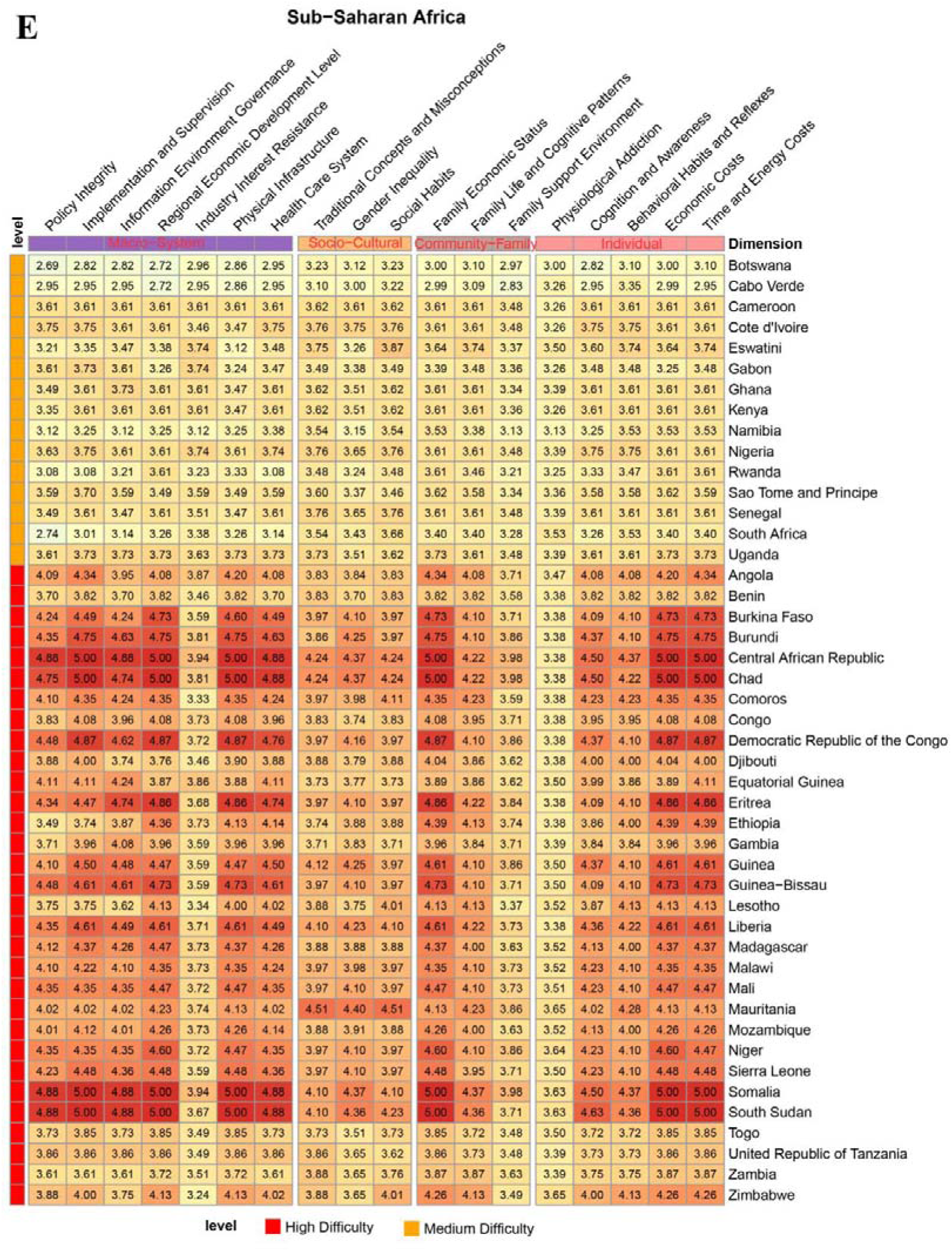
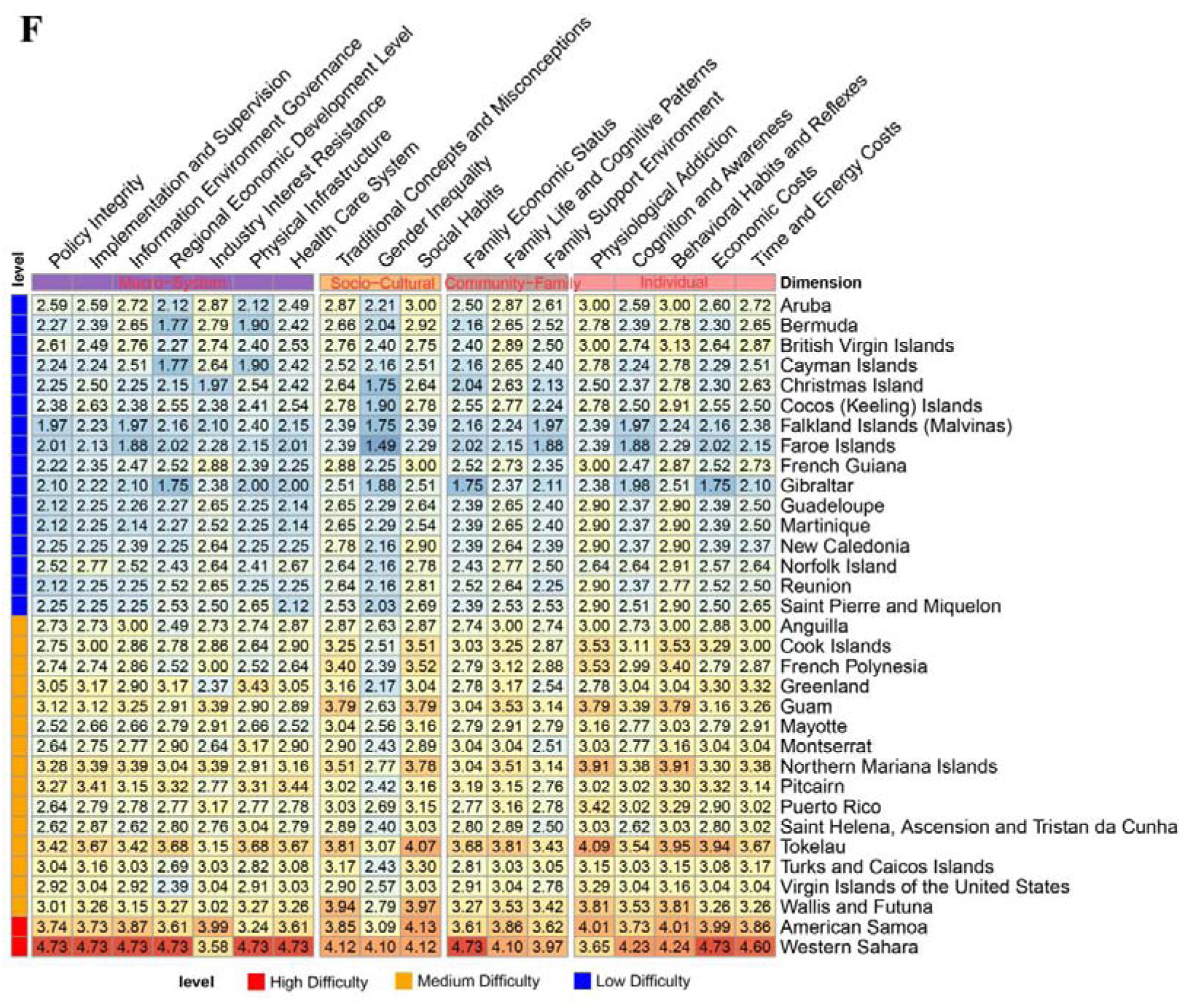
Intervention difficulty scores of high BMI for 18 indicators of countries in (A) Asia, (B) Europe, (C) Latin America, North America, and Caribbean, (D) North Africa and Middle East, Australasia, and Oceania, (E) Sub−Saharan Africa, and (F) others.

On the contrary, countries in North Africa and Middle East achieved the mean intervention score of 3.45, including Kuwait (2.83), Saudi Arabia (2.92), Jordan (3.25), Egypt (3.34), etc. While high-income oil-rich countries like Kuwait and Saudi Arabia offer good private medical care and a high standard of family life, fast food culture and the popularity of traditional sweet treats dominate. Middle-income countries such as Jordan and Egypt, with diets high in oil and sugar, have accessible medical care but lack strong preventive measures (Fig. 3D). Oceania, which has the highest age-standardized prevalence of overweight [6], had a mean intervention score of 3.71, represented by countries like Nauru (4.12), Tonga (3.74), and Samoa (3.51). These nations have middle-income economies but weak medical systems and limited infrastructure, compounded by traditional high-starch diets and imported processed foods. Public awareness of the need for weight management is also low (Fig. 3D).

Sub-Saharan Africa had the highest mean intervention score of 3.88, with countries like Zambia (3.70) and Zimbabwe (3.92) reflecting this trend. These nations have low-to-middle incomes and healthy traditional diets, but there is limited awareness of obesity risks. In urban areas, the consumption of sweetened beverages is widespread, and although obesity is not yet a major public health issue, intervention efforts are hindered once it does occur (Fig. 3E).

### Main factors influencing of unequal difficulty of high BMI intervention

We evaluated the difficulty of high BMI intervention based on 18 indicators from four dimensions, namely “Macro-System Level”, “Socio-Cultural Level”, “Community-Family Level”, and “Individual Level” (Fig. 3, Supplementary part 5, Supplementary Figure 19-36). The SHapley Additive exPlanation (SHAP) method was applied to evaluate the impact of indicators on the unequal intervention difficulty (Fig. 4A). Among them, “Cognition and Awareness” showed the most significant impact (Fig. 4A, Fig. 4B), with the SHAP value of 31.03, followed by “Family life and cognitive patterns” (18.08), “Health Care System” (11.7), “Time and Energy Costs” (11.11), and “Implementation and Supervision” (10.92).

**Figure 4.**
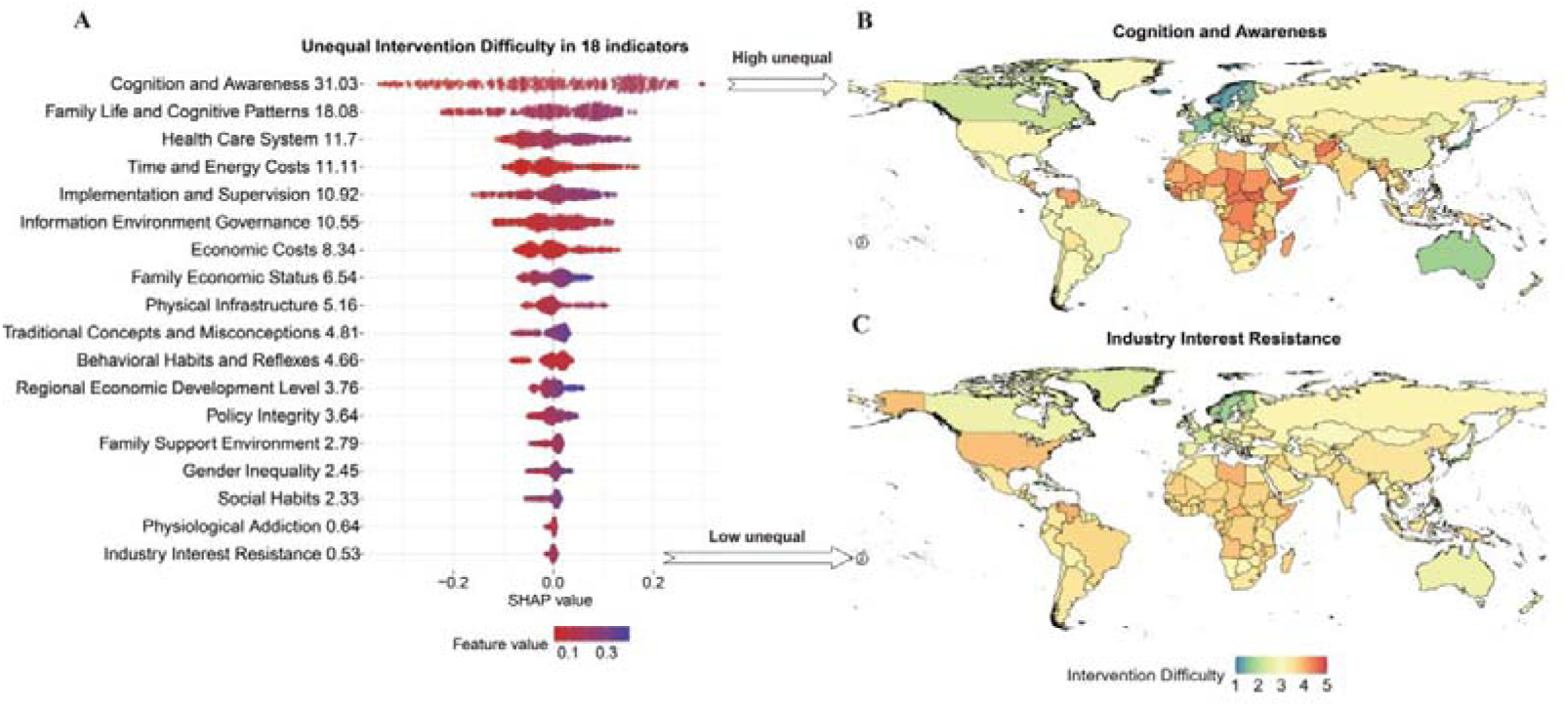
The impact of 18 indicators on unequal difficulty of high BMI intervention. (A) SHAP summary plot of the 18 indicators. (B) The highest unequal indicators, Cognition and Awareness. (C) The lowest unequal indicators, Industry Interest Resistance.

The factors at the “Individual Level” had the greatest influence on intervention difficulty, including “Cognition and Awareness”, “Time and Energy Costs”, “Economic Costs”, and “Behavioral Habits and Reflexes”. “Cognition and Awareness” evaluates an individual’s health literacy, ability to actively learn and apply scientific knowledge, and awareness of their own health risks. Countries like Iceland, Norway, and Sweden had the lowest intervention scores for “Cognition and Awareness” (1.24), while Yemen and South Sudan had the highest scores (4.63), showing a 3.73-fold difference (Fig. 3). “Time and Energy Costs” also significantly impacted the intervention difficulty, with a SHAP value greater than 10 (Fig. 4A). This factor assesses whether individuals have enough time and energy to plan and maintain a healthy lifestyle.

The second most important factors are at the “Macro-System Level”, which encompass political, economic, technological, and public service systems. Among these indicators, the “Health Care System” had the greatest impact. It evaluates whether the system can provide general and preventive weight management and nutrition guidance, as well as treat obesity-related diseases. In cases where early prevention and weight management support are lacking, or where professional resources are scarce and medical services are expensive, the intervention difficulty is significantly higher. “Implementation and Supervision” also had a considerable impact, with a SHAP value of 10.92 (Fig. 4A). This indicator assesses whether regulations are strict, enforcement is efficient, and violations (such as false health claims) are severely punished. Norway had the lowest intervention difficulty for “Implementation and Supervision” (1.24), while countries such as Afghanistan, the Central African Republic, Chad, Somalia, South Sudan, and Yemen had the highest scores (5), showing a 4.03-fold difference (Fig. 3). Another significant indicator was “Information Environment Governance”, which evaluates whether the public information environment is clean, whether unhealthy food advertisements are restricted, and whether scientific health information is easily accessible.

Factors at the “Community-Family Level” also played a significant role in the difficulty of high BMI intervention, including “Family Life and Cognitive Patterns”, “Family Support Environment”, and “Family Economic Status”. These factors assess whether families practice a healthy lifestyle, support healthy health concepts and daily routines, whether family members understand and assist with an individual’s health efforts, and whether a family can afford healthy food, fitness expenses, and professional consulting services. Norway and Denmark had the easiest intervention conditions at this level, while Yemen and Somalia experienced the greatest difficulty, with nearly 3 times the difference in scores (Fig. 3).

Lastly, the factors at the “Socio-Cultural Level” had a relatively smaller impact, including “Traditional Concepts and Misconceptions”, “Gender Inequality”, and “Social Habits”, all with SHAP values less than 5 (Fig. 4A). These factors evaluate the mainstream cultural norms, values, and social atmosphere. “Traditional Concepts and Misconceptions” assesses whether mainstream views support health, whether scientific concepts are widely disseminated, and whether there are common dietary misconceptions. “Gender Inequality” evaluates whether both genders have equal freedom and support in food choices, exercise time, and bodily autonomy. Finally, “Social Habits” reflects whether healthy eating and regular exercise are the societal norms and mainstream social activities.

### Differences in difficulty of high BMI intervention across SDI levels

The difficulty of high BMI intervention is significantly correlated with SDI (R = –0.847, p-value = 1.1×10^-56^, Fig. 5A). A higher the SDI value was associated with lower intervention difficulty.

**Figure 5.**
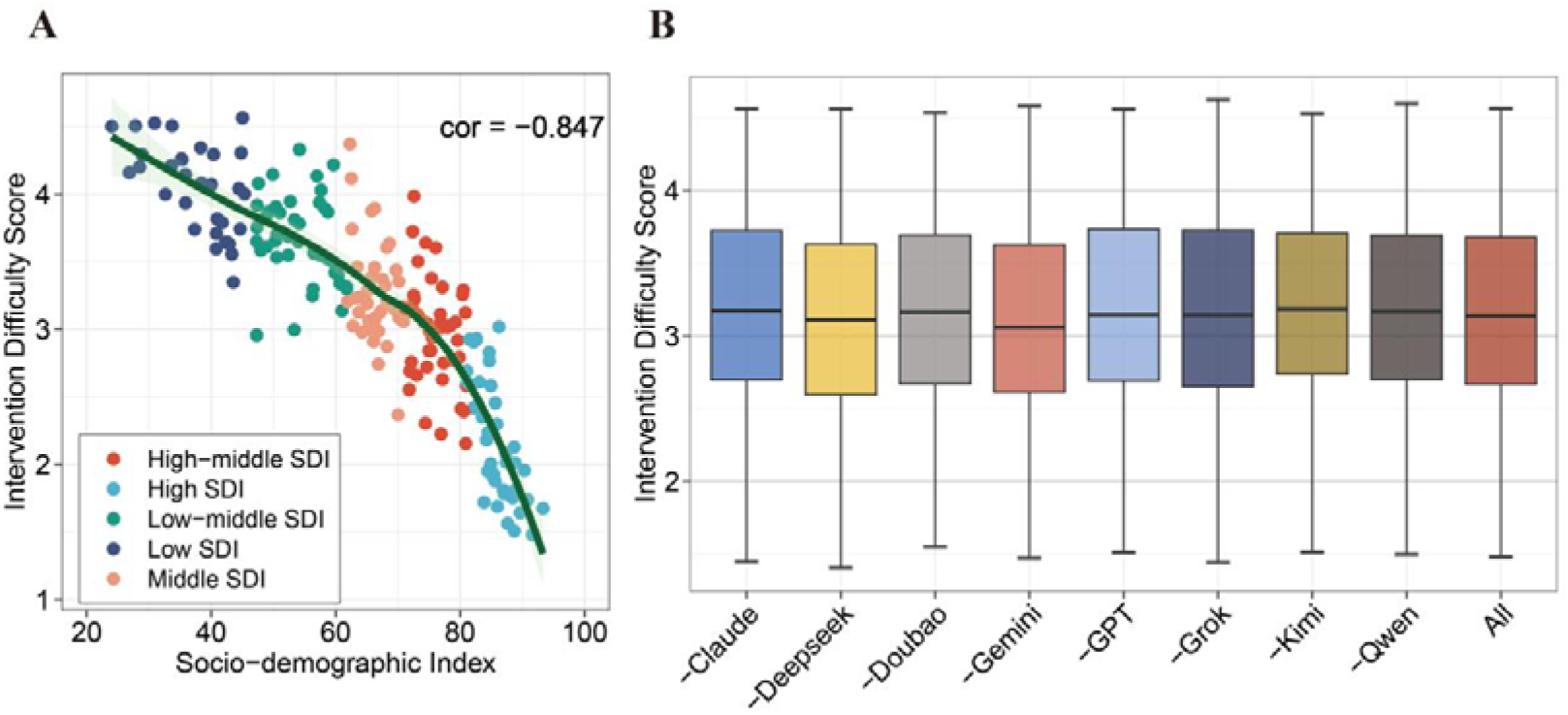
(A) The correlation between intervention difficulty scores of high BMI and SDI values. (B) The sensitivity analyses iteratively excluding each AI model and overall results.

In the 40 locations with a high SDI, the mean intervention difficulty score is 2.18, with Norway having the lowest score and the United States of America being the only country with a score higher than 3. In the 46 locations with a high-middle SDI, the mean intervention difficulty score is 2.98, ranging from the lowest in Israel (2.15) to the highest in Libya (3.99). Notably, 54.35% of countries in this level have scores above 3. In locations with a middle SDI, the mean intervention difficulty score increases to 3.27, with 82.5% of locations having scores above 3. Nauru (4.12) and the Syrian Arab Republic (4.37) have intervention difficulty scores above 4.

In the 42 locations with a low-middle SDI, the mean intervention difficulty score rises to 3.67, with only Bhutan scoring below 3 (2.95), and 6 locations having scores above 4. The highest intervention difficulty is observed in locations with a low SDI, where the mean intervention difficulty score is 4.06, and 64.16% of these locations have scores above 4.

### Independent external empirical verification demonstrated high consistency between intervention difficulty and increase in annual prevalence of obesity, population mean BMI, and national policies

To verify the reliability of results, we compared the GAME intervention difficulty scores against the established baselines, including global increase in annual population mean BMI [31], prevalence of obesity [32], and national policies.

We observed a strong alignment between GAME intervention difficulty and real-world obesity trends (Fig. 6A). Specifically, the 10 countries ranked most difficult to intervene in by the GAME framework exhibited a upward trajectory in the annual increase of age-standardised prevalence of obesity from 1990 to 2022. In contrast, the 10 countries deemed easiest to intervene in showed a declining trend in obesity growth over the same period. Notably, from 1990 to 2002, both groups experienced rising obesity prevalence. The annual increase was even higher in the easier-to-intervene countries during this early phase. However, a marked divergence emerged from 2003 onward. The annual increase in obesity prevalence decelerated in the easier-to-intervene countries, while it continued to rise in the more difficult-to-intervene countries. Under current trends, the annual increase in easier-to-intervene countries is gradually leveling off, whereas it remains on an upward trajectory in more difficult-to-intervene countries. The trends of annual increase of age-standardised prevalence of obesity are highly consistent with GAME intervention difficulty rating.

**Figure 6.**
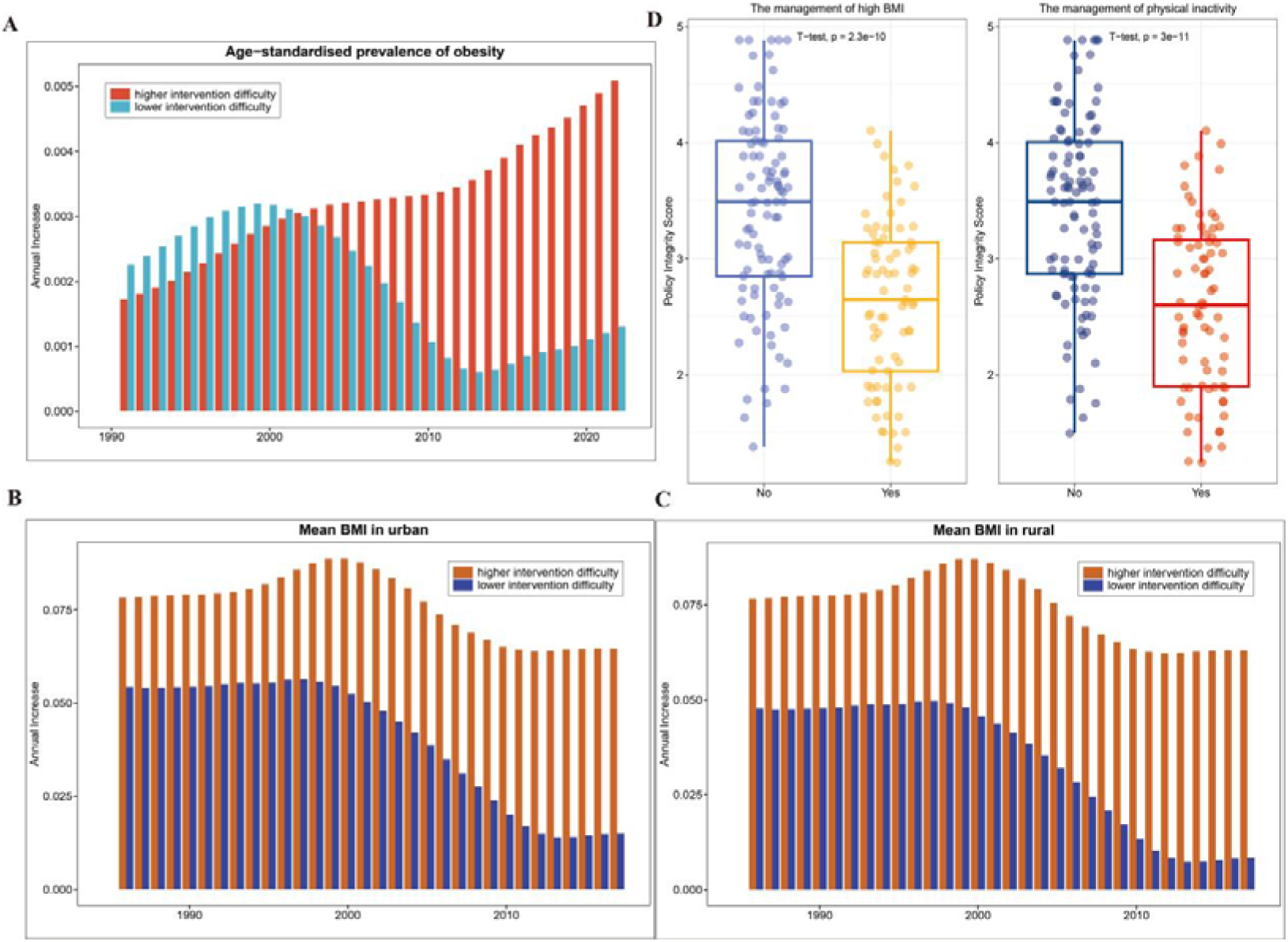
Independent external empirical verification between intervention difficulty scores of high BMI and (A) increase in annual prevalence of obesity, (B) population mean BMI in urban, (C) population mean BMI in rural, and (D) national policies for the management of high BMI and physical inactivity.

The similarly consistent pattern was observed in annual increase of population mean BMI. From 1985 to 2000, urban populations in both easier– and more difficult-to-intervene countries exhibited relatively stable trends in BMI growth (Fig. 6B). However, a clear divergence emerged after 2000. From 2000 to 2017, the annual increase in mean BMI in urban areas declined markedly in easier-to-intervene countries, whereas the decline in more difficult-to-intervene countries was limited. This divergence extended to rural populations as well (Fig. 6C). Moreover, the annual increase in population mean BMI remained consistently higher in more difficult-to-intervene countries than in easier-to-intervene ones from 1985 to 2017, particularly in rural areas. The findings further supported the validity of the GAME framework.

Additionally, we divided countries to two groups according to the presence or absence of national guidelines or standards for the management of high BMI. A highly significant difference was observed in the “Policy Integrity” intervention difficulty scores between the two groups (T-test, p-value = 2.3 × 10^-10^_,_ Fig. 6D). The scores in countries without such guidelines were higher than those with established guidelines. Similarly, the significant difference was also founded when comparing countries with and without national guidelines or standards for the management of physical inactivity (T-test, p-value = 3 × 10^-11^ Fig. 6D). The results also demonstrated that the GAME framework is reliable in assessing the high BMI intervention of different countries.

Together, these findings provided robust and multi-dimensional evidence supporting the GAME framework to characterize global heterogeneity in high BMI intervention challenge.

### Internal evaluation-low heterogeneity and sensitivity of 8 AI

The evaluations of the intervention difficulty across 226 locations, using eight distinct AI models based on 18 key indicators, demonstrated consistently low heterogeneity. For 99.9% indicators, the I² values were below 20% and Cochran’s Q statistic were non-significant (all p > 0.05) (Supplementary Table 2-19). These results indicated a high concordance among the different AI methodologies in ranking global intervention challenges, supporting the reliability of the generated difficulty scores.

Furthermore, sensitivity analyses were performed by sequentially excluding the results from each AI model. The results revealed that the final conclusions remained essentially unchanged regardless of which model was removed, underscoring the robustness of the primary findings (Fig. 5B).

## Methods

### The GAME framework

We evaluated the difficulty of high BMI intervention using the Generative AI Meta-Evaluation (GAME) framework, which is structured across four levels: “Macro-System Level”, “Socio-Cultural Level”, “Community-Family Level”, and “Individual Level”. Each of these levels was further divided into two sub-levels to reflect both their potential impact and measurement difficulty. After conducting a comprehensive manual review of relevant literature in alignment with these dimensions, we identified and selected 18 key indicators that influence intervention difficulty. These indicators are “Policy integrity”, “Implementation and supervision”, “Information environment governance”, “Regional economic development level”, “Industry interest resistance”, “Physical infrastructure”, “Health care system”, “Traditional concepts and misconceptions”, “Social habits”, “Gender inequality”, “Family economic status”, “Family life and cognitive patterns”, “Family support environment”, “Physiological addiction”, “Cognition and awareness”, “Behavioral habits and reflexes”, “Economic costs”, and “Time and energy costs” (Supplementary part 2). Each of these indicators is scored on a scale from 1 to 5, with higher scores representing greater difficulty in implementing interventions. We assigned a weight to each indicator such that the sum of all weights equals 1 (Table 1).

**Table 1.** The 18 indicators in GAME framework.

| Level 1 | Level 2 | Indicators | Weights |
| --- | --- | --- | --- |
| Macro-System Level | Policy and Governance | Policy integrity | 0.0064 |
|  |  | Implementation and supervision | 0.0064 |
|  |  | Information environment governance | 0.0032 |
|  | Economic and Industry | Regional economic development level | 0.084 |
|  |  | Industry interest resistance | 0.056 |
|  | Infrastructure and Public Services | Physical infrastructure | 0.04 |
|  |  | Health care system | 0.06 |
| Socio-Cultural Level | Culture and Beliefs | Traditional concepts and misconceptions | 0.09 |
|  | Social customs and norms | Gender inequality | 0.08 |
|  |  | Social habits | 0.08 |
| Community -Family Level |  | Family economic status | 0.08 |
|  |  | Family life and cognitive patterns | 0.06 |
|  |  | Family support environment | 0.06 |
| Individual Level | Biology and Cognition | Physiological addiction | 0.045 |
|  |  | Cognition and awareness | 0.045 |
|  | Behavior and Cost | Behavioral habits and reflexes | 0.024 |
|  |  | Economic costs | 0.018 |
|  |  | Time and energy costs | 0.018 |

We utilized 8 AI models to assess the intervention difficulty of the 18 indicators for the 226 locations. The AI models used were GPT (https://chatgpt.com/), Deepseek (https://www.deepseek.com/), Doubao (https://www.doubao.com/chat), Grok (https://grok.com/), Qwen (https://www.tongyi.com/), Gemini (https://gemini.google.com/), Claude Sonnet 4 (https://claude.ai/), and Kimi (https://www.kimi.com/) (Supplementary part 1). For each location, we submitted identical questions to all eight AI models separately, asking them to score the 18 indicators on a scale from 1 to 5 (Supplementary parts 2 and 3). The scores from each model were then integrated to produce a combined score for each indicator, using meta-analysis combined with SuperCLUE (https://www.superclueai.com/), a comprehensive evaluation system for general AI. The combined score of each indicator for every country and region is calculated by:

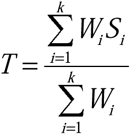

of which, T is the combined score, k is the number of AI, *S_i_* is the score produced by AI_i_, *_Wi_* is the weight of AI_i_ from SuperCLUE:

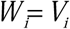

*_Vi_* represents the score of AI_i_ in SuperCLUE.

We tested the heterogeneity of AI models using I^2^ and Cochran’s Q (Supplementary part 4). In addition, we performed a clustering analysis on the scores of the 18 indicators generated by the eight AI models (Supplementary part 4, Supplementary Figure 1-18). Sensitivity analyses were also conducted by iteratively excluding the results from each AI model.

Finally, the intervention difficulty score for each country and region was calculated as the weighted sum of all 18 indicators.

### Statistical analysis

The SDI values for each location were derived from the Global Burden of Diseases, Injuries, and Risk Factors Study (GBD) 2021 (Supplementary Table 20). The SDI is a composite measure that reflects the social and economic conditions influencing health outcomes across different regions. It is calculated as the geometric mean of three indices: the total fertility rate (TFR) for individuals under 25 years old (TFU25), the average education level for individuals aged 15 and older (EDU15+), and per capita lag-distributed income (LDI). In the GBD 2021 study, the SDI values were scaled by multiplying them by 100, resulting in a range from 0 to 100. To measure the correlation between the intervention difficulty scores for high BMI and SDI values, we used Pearson’s correlation coefficient. A significant correlation was defined by an R² value greater than 0.5 and a p-value less than 0.05.

We also employed SHAP values to evaluate the overall importance of various indicators in determining the difficulty of intervening in high BMI. SHAP applies a game-theoretic approach to interpret model behavior by aggregating local feature contributions into global explanations [33, 34]. This methodology is considered more informative than most global approximation techniques. In addition to quantifying the overall indicator importance, SHAP helps to clarify how each individual variable influences specific predictions.

The age-standardised prevalence of obesity from 1990 to 2022 and the population mean BMI in both urban and rural from 1985 to 2017 were obtained in NCD Risk Factor Collaboration [31, 32]. A total of 197 countries were matched to the GAME framework. The annual increase of age-standardised prevalence of obesity was calculated by subtracting the prevalence of the previous year from the current year. The same method was applied to calculate the annual increase of population mean BMI. The existence of national guidelines or standards for the management of high BMI and physical inactivity in 194 countries were obtained from the World Obesity Atlas 2025 [35]. The difference between the countries with and without the guidelines was calculated using the two-sample student T-test.

Data analysis was conducted using the R statistical software package (v4.2.1). For statistical analysis, we used the following R packages: ggbeeswarm, tidyverse, stringr, iml, and randomForest.

## Discussion

Despite global consensus on the severe health threats of overweight and obesity, the prevalence has persistently increased worldwide over the past three decades, affecting all age groups and sexes. High BMI intervention is a leading global health challenge. Here, we developed a GAME framework to evaluate the difficulty of high BMI intervention by integrating 18 indicators from the broad societal structures to the individual’s inner world. Our study revealed the imbalanced intervention difficulty across 226 locations. The results were freely available at http://www.deepburden.com/high-bmi (Supplementary part 6, Supplementary Figure 37-41).

The top 5 easiest locations to intervene in were Norway [1.48], Sweden [1.51], Iceland [1.56], Denmark [1.64], and Switzerland [1.67]. Almost 3 times difference was observed between the easiest locations to intervene in, Norway, and the most difficult locations, Yemen [4.56]. In the 57 locations with an intervention difficulty score lower than 2.67 (the 25th percentile value), 56.14% were classified as High SDI level, followed by 15.79% in the High-middle SDI level, and 1.75% in the Middle SDI level (the other 26.32% had no SDI values). The Middle SDI level locations that demonstrated lower difficulty of high BMI intervention was Costa Rica [2.37]. Notably, five Middle SDI level locations exhibited higher intervention difficulty scores than 3.68 (the 75th percentile value), including Tonga [3.74], Equatorial Guinea [3.88], Iraq [3.89], Nauru [4.12], and Syrian Arab Republic [4.37]. In addition, the findings highlighted that the difficulty of intervening in high BMI is influenced by a variety of factors across different levels. The most significant challenges arise from individual awareness and resources, followed by macro-level systemic factors, while socio-cultural factors play a comparatively smaller role.

We presented the first systematic attempt to evaluate the difficulty of high BMI intervention on a global scale. It offers a novel and comprehensive perspective for understanding multi-level determinants to address the obesity crisis. The GAME framework integrates a wide range of indicators spanning macro-system, socio-cultural, community-family, and individual levels, enabling a holistic view of the factors influencing unequal intervention difficulty. Furthermore, the use of eight leading AI models enhances the robustness of the estimates. Nevertheless, our study has several limitations that should be considered. First, the findings are dependent on the complex AI systems, the interpretability of specific scores for certain locations or indicators may lack transparency. Second, we applied the annual increase of obesity prevalence to verify the reliability of the results. Because the direct evidence of success interventions of high BMI for all 226 locations is constrained, particularly in settings with limited digital infrastructure or inconsistent reporting.

Overall, we provide the first large-scale, AI-driven evaluation of the difficulty of high BMI intervention across 226 locations, offering a crucial evidence to evaluate the effectiveness of interventions and reduce global obesity.

## Ethics approval and consent to participate

Not applicable.

## Consent for publication

Not applicable.

## Availability of data and materials

Not applicable.

## Competing interests

The authors declare no competing interests.

## Funding declaration

This work was supported by the National Natural Science Foundation of China [Grant Nos. 31970651, 92286018]; the Excellent Youth Support plan of Education Department of Heilongjiang Province [Grant No.YQJH2023036]; Marshal Initiative Funding [Grant No. HMUMIF-22010]; XingLian Outstanding Talent Support Program 2024; the Joint Funds of the Zhejiang Provincial Natural Science Foundation of China [Grant No. LBY24H170001].

## Author Contributions

C.S., C.W.L., W,H,L. and W.S. contributed equally to this work. Y.S.J. conceived and contributed the work. C.S., C.W.L., W,H,L., W.S., S.Y.W., H.Y.C., J.X.T., J.C.W., Y.P.Z., L.N.Y., R.L.L., J.X., T.Y.L., Q.W., Y.X., N.W., Y.G., Q.D.R., C.W., S.L.L., C.Y., J.Y.H., T.Y.Z., Q.H.L., Z.W.S, M.M.Z., H.C.L., and Y.S.J. drafted and modified the manuscript.

## Data Availability

All data produced in the present study are available upon reasonable request to the authors

http://www.deepburden.com/high-bmi

## Acknowledgements

Not applicable.

## Notes

### Competing Interest Statement

The authors have declared no competing interest.

### Summary of Updates

Supplemental files updated. We have deleted the redundant image file.

