## Supplementary file for "The Generative AI Meta-Evaluation (GAME) Study Framework: Global, Regional, and Country-Specific Unequal Difficulty of High BMI Intervention"

### Chen Sun et al.

* Correspondence to Yongshuai Jiang, College of Bioinformatics Science and Technology, Harbin Medical University, 194 Xuefu Road, Nangang District, Harbin, Heilongjiang Province, China.

1. **Eight AI models.**

This study uses eight popular AI models (GPT, Claude, Gemini, Grok, Doubao, DeepSeek, Qwen, and Kimi) to score the intervention difficulty score in 226 locations：

1) **GPT-5**

GPT-5 (https://chatgpt.com/) is the fifth-generation generative pre-trained transformer model released by OpenAI. Across multiple benchmarks, GPT-5 is touted as "smarter across the board" in areas such as mathematics, science, law, and programming. OpenAI's official website states that GPT-5 integrates "thinking" capabilities, enabling it to better apply reasoning and strategy to complex tasks. Compared to previous versions, GPT-5 is expected to be even more effective at reducing hallucinations. In the ChatGPT service, users can enhance their interactions by adopting different roles, styles, and tool-based functions.

2) **Grok4**

Grok4 (https://grok.com/) is a model series launched by xAI, an AI project under Elon Musk. Grok 4 is its newest version. In many reviews and blog posts, Grok 4 is considered competitive in reasoning and complex task processing. Grok 4's context window is said to be larger, with some resources claiming support for up to 256k tokens of context input. However, some comparison articles suggest that its performance on coding and programming tasks is not necessarily significantly better than Claude 4 or Opus.

3) **Claude Sonnet 4**

Claude Sonnet 4 (https://claude.ai/): Claude is a series of models released by Anthropic, and "Sonnet 4" is a version of this series. In the SWE-bench (Software Engineering/Programming Benchmark) test, the Claude Sonnet 4 outperformed the Gemini 2.5 Pro. In some tests, Claude has received industry recognition for its reasoning, multitasking capabilities, and reliability. Claude's design emphasizes security, alignment, and controllability.

4) **Gemini 2.5 Pro**

Gemini 2.5 Pro (https://gemini.google.com/): Gemini is a family of AI products from Google/DeepMind, designed to be a multimodal, tool-enabled, and universal intelligent assistant. "2.5 Pro" is an advanced version/variant of Gemini, focusing on reasoning and tool usage capabilities. In some evaluations, Gemini 2.5 Pro performed well in tasks such as visualization, data processing, and information retrieval. In one evaluation, Gemini 2.5 Pro (along with DeepSeek) was ranked as one of the models with low token usage efficiency (i.e., low cost for a given task).

5) **Doubao-Seed-1.6-thinking**

Doubao-Seed-1.6-thinking (https://www.doubao.com/chat): Doubao is an AI chat/large-scale model project under ByteDance/Volcengine. "Doubao 1.6," a newer version, is advertised as supporting an "adjustable thinking depth" mechanism. In the "LobeChat" model information, Doubao-Seed-1.6-Thinking is described as a version with "enhanced reasoning capabilities." LobeHub media reports indicate that Doubao-1.6 introduces upgrades such as multimodal and tool-calling capabilities. For example, "Doubao 1.6-Vision" supports image processing/tool-calling for visual reasoning and image manipulation tasks (cropping, scaling, rotation, and annotation). Users and the system can select different "thinking depth" modes, such as Minimum, Low, Medium, and High, based on task complexity, to achieve a balance between efficiency and quality. In the AIBase report, it was also stated that Doubao-1.6 introduced a lite version to adapt to enterprise scenarios and reduce usage costs.

6) **Kimi-K2**

Kimi-K2 (https://www.kimi.com/): Kimi K2 is a newer model released by Moonshot AI, an upgrade to its Kimi series. On Hugging Face's model page, Kimi K2 (Instruction version) is described as trained using the Muon optimizer, boasting excellent performance on cutting-edge knowledge, reasoning, and coding tasks, and optimized agentic capabilities (autonomous invocation, task decomposition, and tool usage). According to Moonshot's official and community resources, Kimi K2 is a Mixture-of-Experts (MoE) architecture model with a total parameter size of 1 trillion, but only a subset of these parameters (32B active parameters) are activated for computation during each inference.

7) **Deepseek-V3.1**

Deepseek-V3.1 (https://www.deepseek.com/): DeepSeek is a model and platform that has emerged in the Chinese and global AI ecosystems in recent years, with its V3 and R1 series widely discussed. DeepSeek V3.1 is an upgraded version of V3, and some reviews and blogs differentiate between "DeepSeek V3.1" and "DeepSeek-3.1 Thinking/Non-thinking" models. An article comparing DeepSeek V3.1 with Kimi K2 mentions that DeepSeek V3.1 introduces a "thinking/non-thinking" mode switching mechanism, using special tokens to enable the model to enter a deeper reasoning/thinking state in certain situations. The article also notes that DeepSeek V3.1's "thinking mode" significantly improves performance over its "non-thinking mode" in mathematical, coding, and long-context reasoning tasks.

8) **Qwen 3**

Qwen 3 (https://www.tongyi.com/): Qwen is a series of large-scale language and multimodal models launched by Alibaba. Qwen 3 is its latest version. Qwen 3 has multiple variants, including dense and sparse models with activation parameters. Multiple public articles and reviews claim that Qwen 3 is one of the strongest open-source models currently available, surpassing DeepSeek-R1 or competing models on certain benchmarks. Some reports indicate that Qwen 3 has surpassed Kimi-2 on certain tasks.

2. **Scoring rules.**

The same questions were asked to these eight AIs to generate 18 scores on the difficulty of quitting smoking in 226 locations. The detailed scoring rules for the 18 indicators are as follows:

1) **Policy integrity: The comprehensiveness and scientific nature of relevant laws and regulations.**

**5 points (Non-Existent)**: There are virtually no targeted laws, regulations, or policy documents in this area. The issue is left unchecked, or there are only ineffective and meaningless statements.

**4 points (Rudimentary)**: The government has begun to recognize the problem and has issued some fragmented, framework-based policy documents. However, these documents are too principled and lack specific implementation details, responsible parties, and punitive measures. This leads to poor operability and very limited coverage.

**3 points (Basic Framework)**: A foundational legal and regulatory framework has been established, covering key aspects of this sector and stipulating basic rights, obligations, and penalties. However, there are significant policy loopholes, or standards differ significantly from international best practices, or policies in different regions are fragmented and inconsistent.

**2 points (Advanced)**: A comprehensive and systematic legal and regulatory framework has been established, covering most key links and scenarios. The policy content is scientific, drawing on successful international practices, and provides clear enforcement entities, strict penalties, and effective implementation mechanisms. However, some minor shortcomings or details still need to be optimized.

**1 point (Comprehensive and Leading)**: The policy system is not only comprehensive and comprehensive, but also leads the world in scientific research, foresight, and innovation. Policy designs not only address current issues but also foresee and mitigate future risks. A comprehensive evaluation and regular revision mechanism has been established to ensure that policies remain optimized. A solid legal foundation and cultural atmosphere have been established throughout society.

2) **Implementation and supervision: The ability and efficiency of policy implementation, including supervision, law enforcement, and punishment mechanisms.** It refers to the actual implementation of promulgated policies and regulations. It examines whether the supervisory system is effective, whether law enforcement is strict and regular, whether penalties are sufficiently deterrent, and the efficiency of the entire implementation process.

**5 points (Lawless)**: The law is virtually nonexistent. There is no effective oversight or enforcement mechanism. Violations are open, widespread, and without consequences. The public has no trust in the law.

**4 points (Symbolic Enforcement)**: Campaign-style, selective enforcement is only carried out on rare, specific occasions, or against unlucky individuals. Enforcement is conducted for show, not to truly uphold the dignity of the law. Violations remain the norm.

**3 points (Partially Effective)**: Law enforcement is effective in some areas, regions, or time periods. However, there are significant gaps in enforcement, regional disparities, and insufficient resources. Violations remain at a high risk.

**2 points (Strict and Effective)**: Law enforcement is regular, strict, and predictable. We have an efficient monitoring and reporting system and a professional law enforcement team. The vast majority of violations are promptly detected and punished harshly and consistently, making the cost of violations very high.

Characteristics: A robust monitoring system (e.g., regular inspections, electronic surveillance, and public reporting); swift law enforcement response; severe and impartial penalties; and high-risk behaviors with a high probability of being caught and high costs.

**1 point (Rule of Law)**: The authority of the law is absolutely respected. Law enforcement is not just a government activity; it is internalized into a shared social culture and a norm to be consciously observed. Violations are extremely rare, and when they do occur, the punishment mechanism operates with exceptional efficiency, ensuring that all violations are brought to justice.

3) **Information environment governance: Regulation of public information, such as regulating advertising and combating false information.** It refers to the ability of governments and relevant institutions to regulate and guide the public information ecosystem, primarily through regulating commercial advertising (such as prohibiting misleading advertising), combating false and misleading information, and promoting scientific knowledge. Its goal is to ensure that the public has access to accurate, scientific, and health-promoting information.

**5 points (Misinformation Rampant)**: The information environment is completely out of control. False information, pseudoscience, and harmful health rumors spread massively and unimpeded through all media, especially social media. Official scientific voices are weak or absent, and commercial advertisements engage in unbridled exaggeration and deceptive propaganda, without any effective oversight.

**4 points (Lax Regulation)**: Some fundamental regulations exist, but enforcement is extremely weak, rendering them virtually ineffective. Commercial advertising often skirts the line, using obscure yet misleading language. Rumors continue to spread widely, and official debunking efforts are slow, sporadic, and limited in impact, failing to effectively mitigate them.

**3 points (Moderate Control)**: The regulatory system is beginning to function, capable of addressing even the most blatant forms of false advertising and information. However, there are significant loopholes and lags. Businesses will continually seek new ways to circumvent regulations. Rumor-debunking mechanisms have been established, but their reach and reach are often inferior to the rumors themselves, resulting in limited effectiveness.

**2 points (Effective Governance)**: A proactive, systematic regulatory system has been established. Regulations are strict and enforcement is strong, enabling rapid response to new marketing tactics and communication channels. Scientific information is disseminated proactively, extensively, and creatively, effectively reaching target audiences and combating rumors. Commercial advertising is strictly limited to facts.

**1 point (High-Quality Info Ecosystem)**: The information environment is ideal. Scientific, evidence-based information has become the absolute mainstream and deeply rooted in public opinion. The public generally possesses high media literacy and scientific discernment, actively resisting and criticizing false information. Illegal commercial advertising and large-scale rumor-mongering incidents have become extremely rare. The focus of governance has shifted from "crackdown" to "maintaining" and "improving".

4) **Regional economic development level: the basis for determining public resource investment and individual payment capacity.** This score assesses a region's overall economic prosperity. It directly determines two aspects, including a) Public resource investment: whether the government has sufficient fiscal revenue to invest in public services and infrastructure, such as healthcare, education, and the environment and b) Personal affordability: whether households have sufficient disposable income to cover health-improving expenses, such as healthy food, medical services, and smoking cessation medications.

**5 points (Extreme Poverty)**: The economy is in a state of collapse or stagnation. Infrastructure is deficient, the government is dysfunctional, and the ability to provide basic public services is inadequate. The vast majority of the population engages in subsistence farming or the informal economy, living below the international poverty line, with survival as the primary concern.

**4 points (Developing and Poor)**: The economy has begun to develop, but the level remains very low. Limited government revenue allows only the most basic public services, which are inadequate in quality and coverage. A high proportion of the population lives in absolute poverty, with a large number of people barely able to meet basic needs and extremely vulnerable to risks (such as illness).

**3 points (Middle-Income)**: The economy is growing rapidly, and the country or region is in the process of industrialization. The government has stable and growing fiscal revenues, enabling large-scale investments in infrastructure and public services (such as universal health care). Absolute poverty has been significantly reduced, and a sizable middle class has emerged, but the gap between rich and poor may be large.

**2 points (Developed)**: The economy is highly developed and entering the post-industrial era, dominated by service and high-tech industries. The government has strong financial resources and is able to provide high-standard, extensive, and high-quality public services and social welfare systems. Residents are generally affluent, with high disposable incomes, and health expenditures account for a reasonable proportion of total expenditures.

**1 point (Highly Developed)**: A global economic leader with exceptionally high levels of national wealth and social welfare. The government invests heavily in public health and social services, striving for ultimate efficiency and fairness. Health is viewed not only as a necessity but also as a fundamental right and a symbol of quality of life.

5) **Industry interest resistance: lobbying and resistance to public policies by vested interest groups.**

**5 points (Extremely strong resistance)**: This industry is a pillar of the national economy and enjoys a unique status of "government-enterprise integration," allowing it to directly veto or significantly weaken policies and regulations that are unfavorable to its products. Public health policymaking is subject to strong interference from this industry.

**4 points (Strong resistance)**: Industries influence legislators and public opinion through powerful lobbying groups, political donations, and advertising spending, which can delay the introduction of effective policies for a long time or water them down.

**3 points (Moderate resistance)**: Industry lobbying and public relations efforts are countered by health advocates. Policy is eventually adopted, but likely as a compromise (e.g., implementation date is delayed, standards are lower than scientific recommendations).

**2 points (Weak resistance)**: The industry's influence has waned, and its opposition has struggled to influence mainstream public opinion and the policy-making process. The main resistance to policy implementation comes from technical details rather than direct industry intervention.

**1 point(No resistance)**: The industry does not exist or has been completely transformed, and public policy is made entirely based on scientific evidence and the public interest, without interference from commercial interests.

6) **Physical infrastructure: basic support such as exercise spaces, electricity, and network.**

**5 points (Extreme Deprivation)**: Severely deficient infrastructure: no safe exercise spaces; even walking is difficult; basic amenities are hard to access.

**4 points (Lack)**: Poor infrastructure: lack of public activity spaces; unsafe or inconvenient environments hinder outdoor activity.

**3 points (Basic Security)**: Adequate infrastructure: cities are car-centric, but basic sports venues are available.

**2 points (Relatively well-developed)**: Good infrastructure: basic recreational spaces exist, but convenience can be improved.

**1 point (Complete and Convenient)**: Excellent infrastructure: safe, connected walking/cycling paths; abundant parks; accessible fitness facilities.

7) **Health care system: access, quality, and equity of healthcare resources.**

**5 points (Inaccessible)**: Basic health care facilities and professional medical personnel are lacking. People cannot access basic care, vaccinations, and emergency services. Medical expenses are entirely out of pocket, and impoverishment due to illness is extremely common.

**4 points (Poor accessibility)**: While the country has a basic healthcare system, resources are extremely strained, waiting times are long, and there are shortages of equipment and medicines. Quality varies, equity is poor, and rural and poor populations face difficulties accessing healthcare.

**3 points (Basic access)**: A universal healthcare network (e.g., the medical insurance system) has been established, addressing most common illnesses. However, high-quality resources are concentrated in major cities, and significant disparities in quality and accessibility remain between different regions and social classes.

**2 points (Good Access)**: The healthcare system is well-developed, and residents have easy access to high-quality, affordable healthcare. Disparities between different population groups are minimal.

**1 point (Accessibility, Quality, and Quality)**: The system is not only efficient and equitable, but also prioritizes prevention and holistic health management. With minimal waiting times and access to world-class medical technology and services, health equity is maximized.

8) **Traditional concepts and misconceptions: widespread social cognitive misconceptions.**

**5 points (Deeply ingrained)**: False beliefs are accepted as truth by the vast majority of people, while scientific knowledge is considered heresy. Efforts to change beliefs are almost ineffective. For example, obesity is either highly stigmatized or completely normalized and weight-loss efforts are mocked or suppressed.

**4 points (Prevalent)**: Misconceptions (e.g., “weight loss means cutting carbs,” “detox with oil”) are widespread in society, hindering the dissemination of scientific knowledge. Traditional ideas dominate everyday decision-making.

**3 points (Coexistence with Science)**: Traditional misconceptions coexist with scientific knowledge, and different groups hold different views. Science communication is effective but has not yet become a social consensus.

**2 points (Occasionally)**: Science becomes mainstream, and misconceptions exist only in a few marginalized groups or specific situations.

**1 point (No misunderstandings)**: Scientific concepts have become a consensus in the whole society, and traditional misunderstandings have basically disappeared.

9) **Gender inequality: Gender stereotypes and power structures in society and culture.**

**5 points (Severe inequality)**: Women have extremely low status and little access to education, work, or independent health decisions. Gender-based violence is widespread.

**4 points (Inequality)**: There is obvious gender discrimination, and women are at a disadvantage in terms of health, education, and access to economic resources, and have limited decision-making power.

**3 points (Moderate Equality)**: Legal equality exists, but in practice, there are "glass ceilings" and hidden social norms. Women have some autonomy in their health decisions, but are still influenced by traditional role expectations.

**2 points (Substantial Equality)**: Gender equality is nearly achieved, with only minor gaps in a few areas. Women have a high degree of autonomy in their health choices.

**1 point (Perfect Equality)**: Gender no longer plays a role in influencing individuals' opportunities, resources, and health choices. Both sexes have equal freedom and support in food choices, exercise time, and bodily autonomy.

10) **Social habits.**

**5 points (Strong Impediment)**: Social activities heavily rely on unhealthy behaviors, and not participating leads to ostracism. For example, high-calorie eating is the only form of entertainment or socializing and healthy behavior is seen as strange..

**4 points (Impediment)**: Unhealthy behaviors are common social tools, with high participation, but a slightly higher tolerance for refusal.

**3 points (Neutral)**: Socializing often involves heavy eating or drinking, but individuals can choose whether to participate.

**2 points (Promote)**: Social habits are generally healthy, though occasional indulgent meals occur.

**1 point (Strong Promoter)**: Healthy eating and regular exercise are social norms and mainstream ways to socialize.

11) **Family economic status: Family income level.**

**5 points (Poverty)**: Family cannot afford any extra health expenses; survival depends on the cheapest calorie sources.

**4 points (Financially Limited)**: Healthy food is a significant cost; reliance on cheap processed foods is common.

**3 points (Subsistence)**: Income meets basic needs for food, must weigh health food against other expenses and premium health products are occasional.

**2 points (Moderately Prosperous)**: Sufficient income allows for easy access to healthy food, fitness, regular checkups, and commercial health insurance. Health spending is stress-free; premium healthy ingredients are prioritized.

**1 point (Affluent)**: Adequate income allows for the freedom to choose any high-quality health services and products, with money no longer a limiting factor in health decisions.

12) **Family life and cognitive patterns: Family health concepts.**

**5 points (Totally Disagree)**: The family holds misguided health beliefs and actively encourages unhealthy behaviors (e.g., feeding children junk food or believing that being overweight is a blessing).

**4 points (Disagree)**: The family is unconcerned about health, adopts a laissez-faire attitude, and has unhealthy eating and lifestyle habits, such as long-term high-fat or high-salt diet, heavy reliance on processed foods, and almost no physical activity.

**3 points (Neutral)**: The family recognizes the importance of health but lacks scientific knowledge or compromises for convenience and taste.

**2 points (Support)**: The family has a good understanding of health and strives to practice a healthy diet and lifestyle, although adherence may be challenging at times.

**1 point (Totally Support)**: Healthy living is a core family value, and the family consistently makes the healthiest choices and continues to learn new things.

13) **Family support environment: emotional, intellectual, and behavioral support provided by family members.**

**5 points (No Support)**: Family actively obstructs (e.g., constantly offers food, buys junk food, criticizes efforts) and creates emotional pressure.

**4 points (Weak Support)**: Family members verbally support the individual but fail to take concrete action, or occasionally act in a counterproductive manner.

**3 points (Moderate Support)**: Family members express understanding and offer some assistance when requested.

**2 points (Good Support)**: Family members actively cooperate and work together to create a healthy environment.

**1 point (Full Support)**: Family members are strong advocates for health, providing emotional encouragement, knowledge sharing, and ongoing behavioral support to help overcome challenges together.

14) **Physiological addiction: Physical dependence on a substance.**

**5 points (Highly Addictive)**: Severe food addiction; eating is out of control; resembles substance addiction with binge-eating behaviors.

**4 points (Moderately Addictive)**: Strong dependency on specific foods (e.g., sweets, carbs); withdrawal causes physical or psychological discomfort.

**3 points (Moderately Addictive)**: There is mild dependence, with significant discomfort during withdrawal.

**2 points (Weakly Addictive)**: There is almost no physical dependence, and it is more of a psychological habit.

**1 point (Non-Addictive)**: The behavior does not involve an addictive substance.

15) **Cognition and awareness: Lack of scientific knowledge, cognitive bias.**

**5 points (Very Poor Cognition)**: Complete lack of relevant knowledge, widespread misconceptions, and a reluctance to accept new information.

**4 points (Poor Cognition)**: Knowledge is fragmented, with major cognitive biases (such as optimism bias) and susceptibility to rumors.

**3 points (Average Cognition)**: Basic common sense is present, but understanding is shallow and difficult to apply to complex decision-making.

**2 points (Good Cognition)**: Comprehensive health knowledge is mastered and can guide most behaviors based on this knowledge.

**1 point (Excellent Cognition)**: Expert-level health literacy is demonstrated, enabling critical evaluation of health information and optimal decision-making.

16) **Behavioral habits and reflexes: automatic behaviors that are conditioned reflexes.**

**5 points (Strong Habit)**: Life is driven entirely by powerful unhealthy habits; nearly all daily choices contradict health goals.

**4 points (Strong Habit)**: The behavior is a strong habit (e.g., dessert after every meal, late-night snacking while scrolling), but can occasionally be controlled with conscious effort.

**3 points (Moderate Habit)**: Old and new habits coexist and conscious effort required to maintain healthy behaviors.

**2 points (Weak Habit)**: Only minor reminders needed to stay on track.

**1 point (No Unhealthy Habit)**: Healthy behaviors are automatic (e.g., post-meal walk, choosing water) and no willpower needed.

17) **Economic costs: Direct costs of adopting healthy behaviors.**

**5 points (Extremely Costly)**: The cost of the health behavior is prohibitive and consumes a significant portion of income.

**4 points (Highly Costly)**: The cost is high and requires careful consideration and savings to afford (e.g., expensive gym personal training).

**3 points (Moderately Costly)**: The cost is affordable, but still an expense that needs to be considered (e.g., the price premium of high-quality health foods over regular foods).

**2 points (Low Cost)**: The cost is very low and barely impacts the household budget (e.g., basic fitness equipment).

**1 point (No Cost)**: The behavior itself incurs no additional cost and may even save money (e.g., taking a walk).

18) **Time and energy costs: The investment required to change behavior.**

**5 points (Extreme Cost)**: Survival stress has drained all mental bandwidth and no cognitive or temporal resources left for any intentional change.

**4 points (High Cost)**: Work or life pressure is overwhelming; carving out time and energy for weight loss is exhausting and unsustainable.

**3 points (Moderate Cost)**: Must sacrifice rest or leisure time; feels somewhat fatigued.

**2 points (Low Cost)**: Busy, but uses efficient time management to maintain health commitments.

**1 point (No Cost)**: Flexible schedule; ample time and energy to plan and execute health plans.

3. **Questioning method**

Giving the scoring rules, the same questions were asked to each of the eight AI models to generate scores across 18 indicators on the difficulty of high BMI intervention in 226 locations: “Based on Policy integrity, Implementation and supervision, Information environment governance, Regional economic development level, Industry interest resistance, Physical infrastructure, Health care system, Traditional concepts and misconceptions, Gender inequality, Social habits, Family economic status, Family life and cognitive patterns, Family support environment, Physiological addiction, Cognition and awareness, Behavioral habits and reflexes, Economic costs, and Time and energy costs, score the difficulty of high BMI intervention in [list of 10 locations per query], and give detailed reasons”. Through repeated queries, we collected scores and corresponding explanatory reasoning for each indicators across 226 locations.

4. **The low heterogeneity of AI models.**

Cochran's Q statistics and I^2^ were employed to evaluate the heterogeneity of AI models:

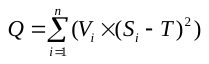

of which,
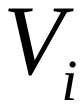
 is the score of AI_i_ in SuperCLUE,
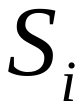
 is the score from AI_i_, and
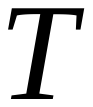
 is the combined score. The p-value for the Cochran's Q statistic is obtained through chi-square distribution.

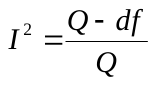

where
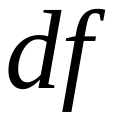
 is the degree of freedom, namely the number of AI is reduced by 1.

Beyond I^2^ and Cochran's Q, we applied the clustering analysis to test the heterogeneity of AI models. The results suggested that there was no uniform pattern for the 8 AI systems about the 18 scoring indicators (Figure S1-S18), thereby enhancing confidence in the generalizability and reproducibility of the intervention difficulty rankings presented in this study.

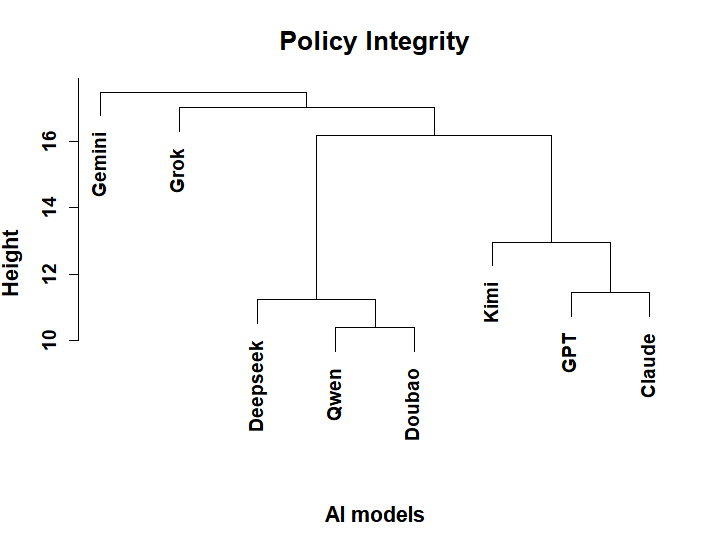

**Figure S1: The clustering analysis of Policy integrity scores in 8 AI models.**

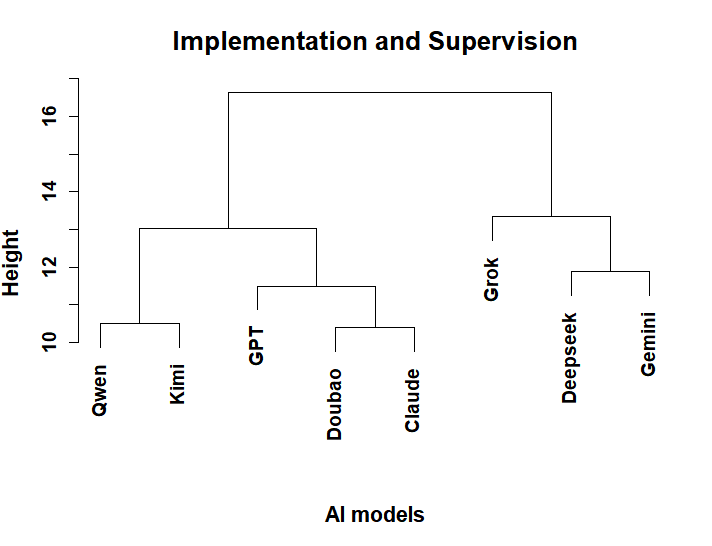

**Figure S2. The clustering analysis of Implementation and supervision scores in 8 AI models.**

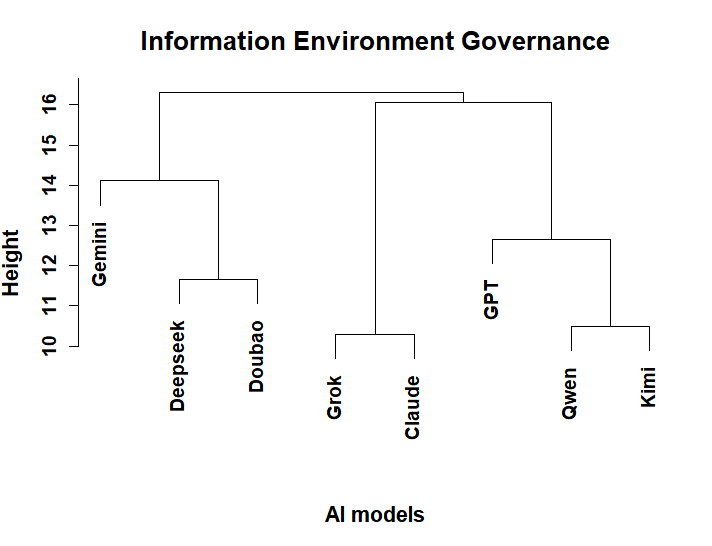

**Figure S3. The clustering analysis of Information environment governance scores in 8 AI models.**

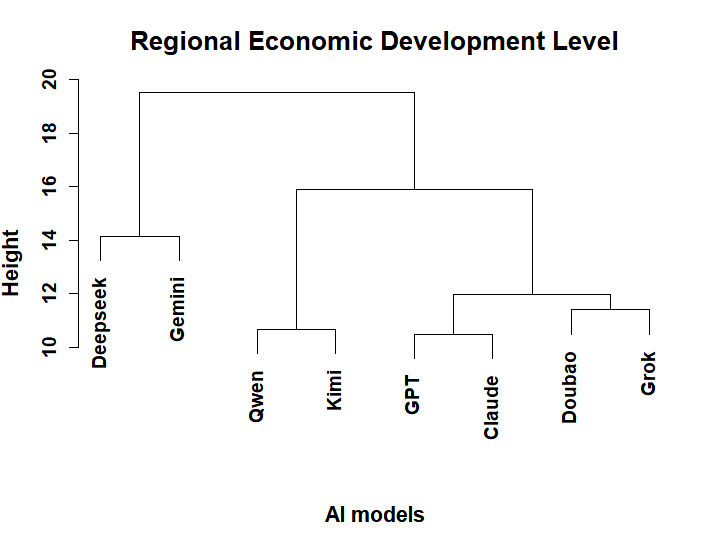

**Figure S4. The clustering analysis of Regional economic development level scores in 8 AI model**
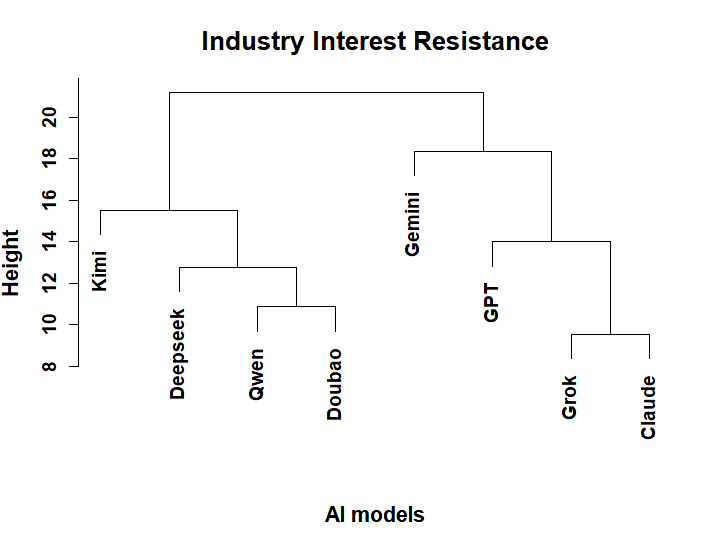

**Figure S5. The clustering analysis of Industry interest resistance scores in 8 AI model.**

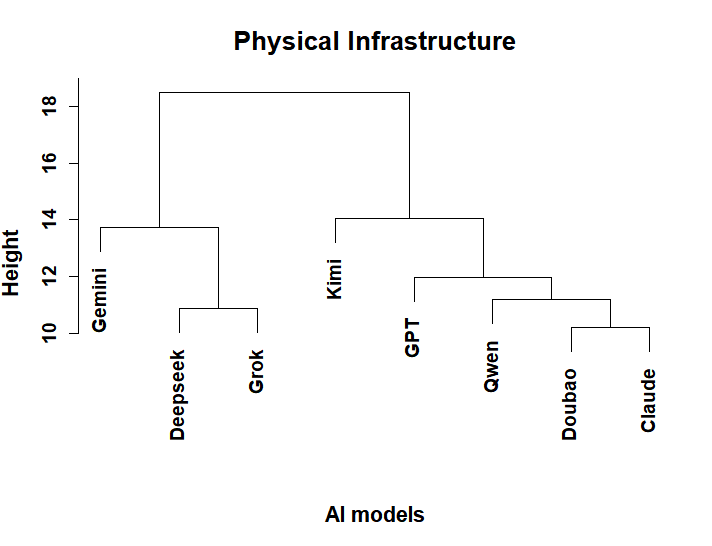

**Figure S6. The clustering analysis of Physical infrastructure scores in 8 AI models.**

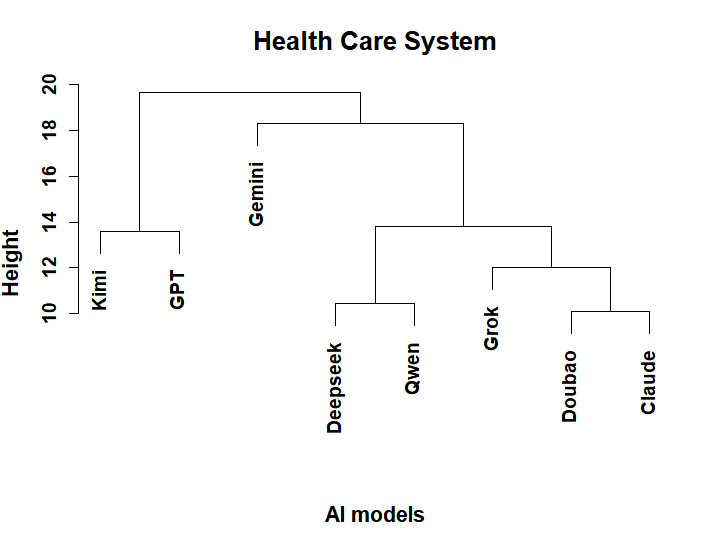

**Figure S7. The clustering analysis of Health care system scores in 8 AI models.**

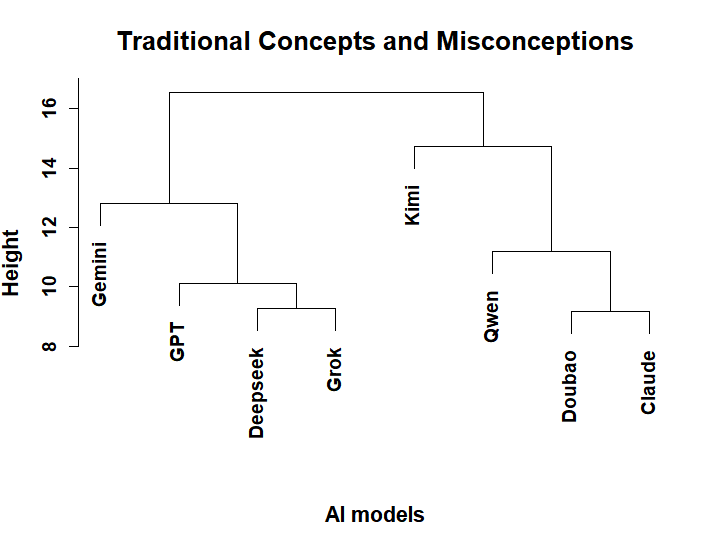

**Figure S8. The clustering analysis of Traditional concepts and misconceptions scores in 8 AI models.**

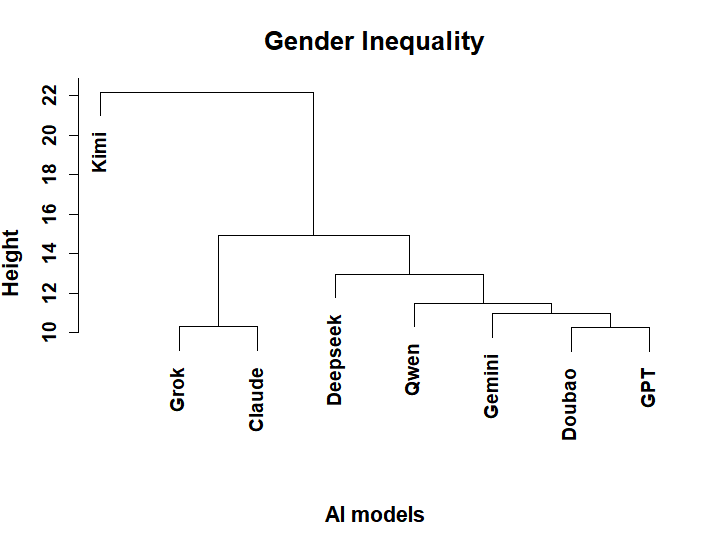

**Figure S9. The clustering analysis of Gender inequality scores in 8 AI models.**

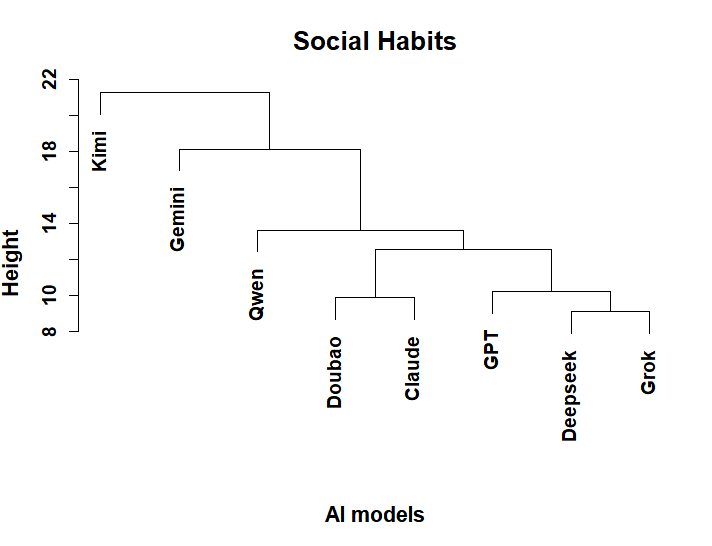

**Figure S10. The clustering analysis of Social habits scores in 8 AI model**

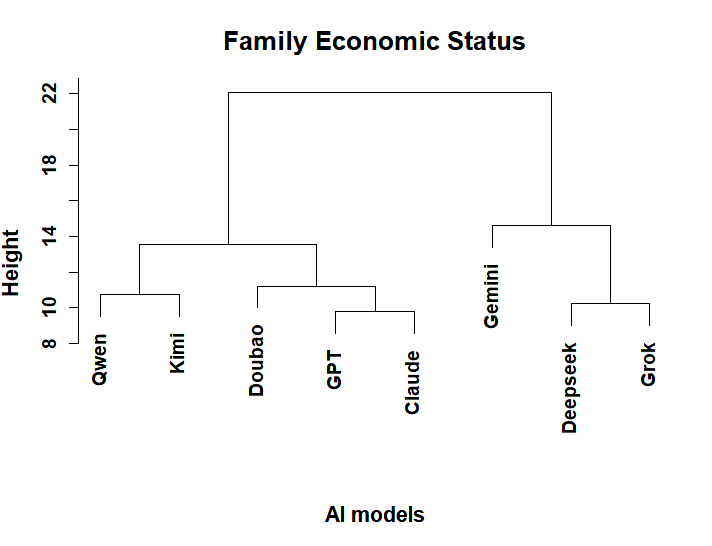

**Figure S11. The clustering analysis of Family economic status scores in 8 AI models.**

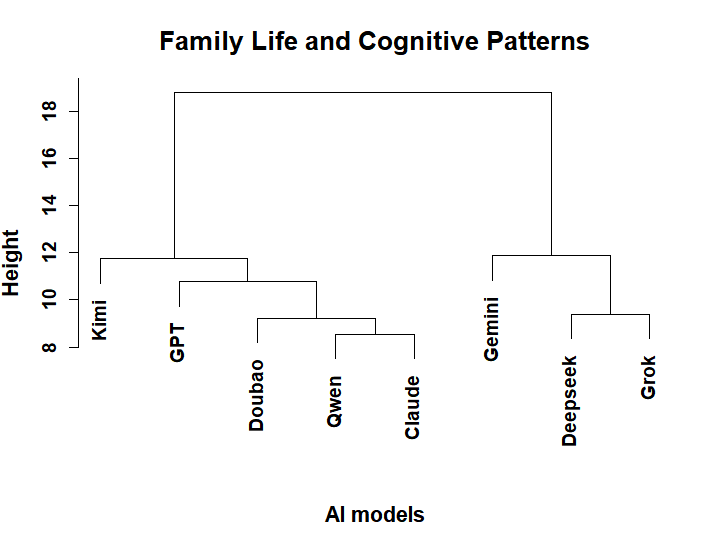

**Figure S12. The clustering analysis of Family life and cognitive patterns scores in 8 AI models.**

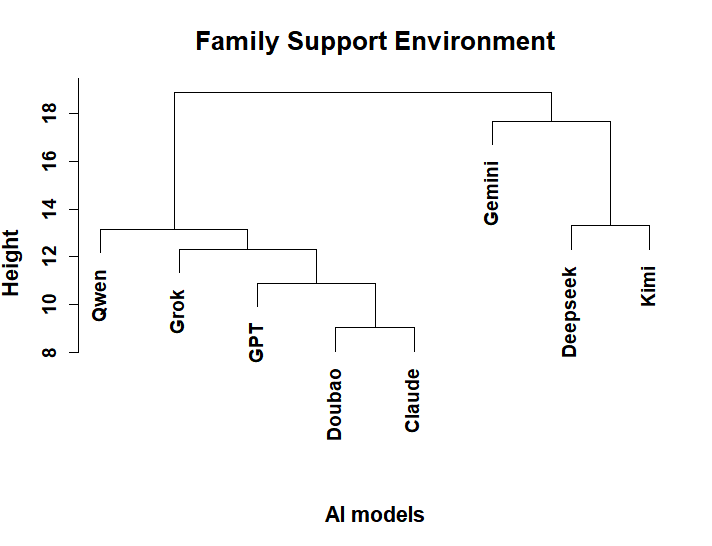

**Figure S13. The clustering analysis of Family support environment scores in 8 AI models.**

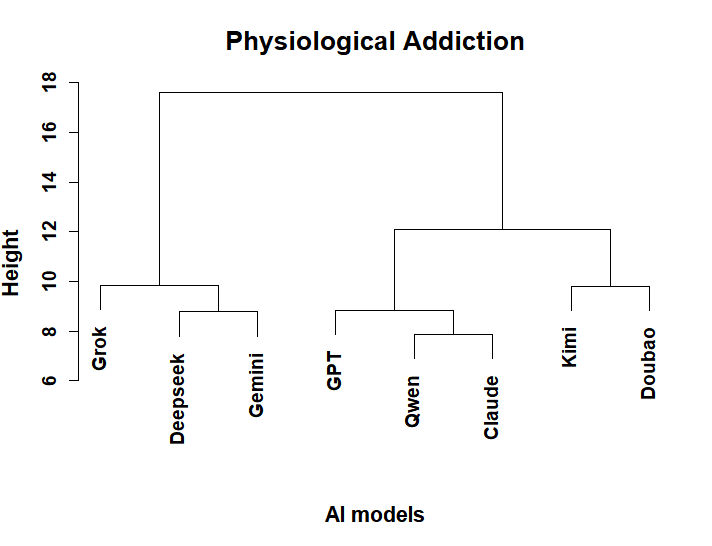

**Figure S14. The clustering analysis of Physiological addiction scores in 8 AI models.**

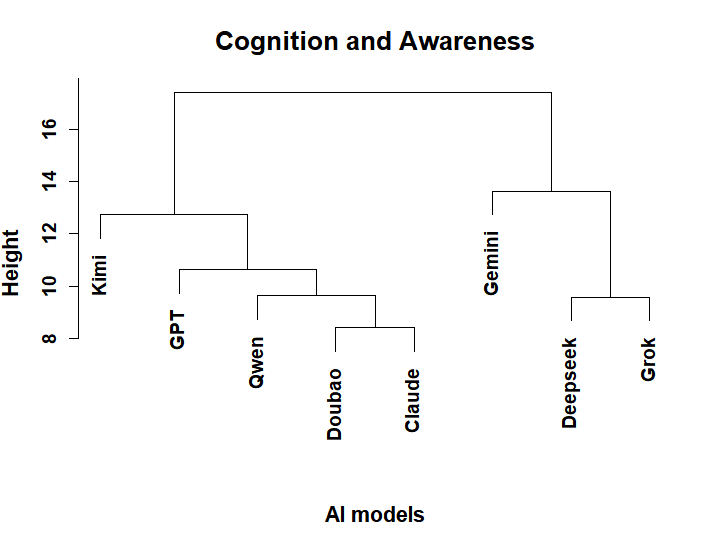

**Figure S15. The clustering analysis of Cognition and awareness scores in 8 AI models.**

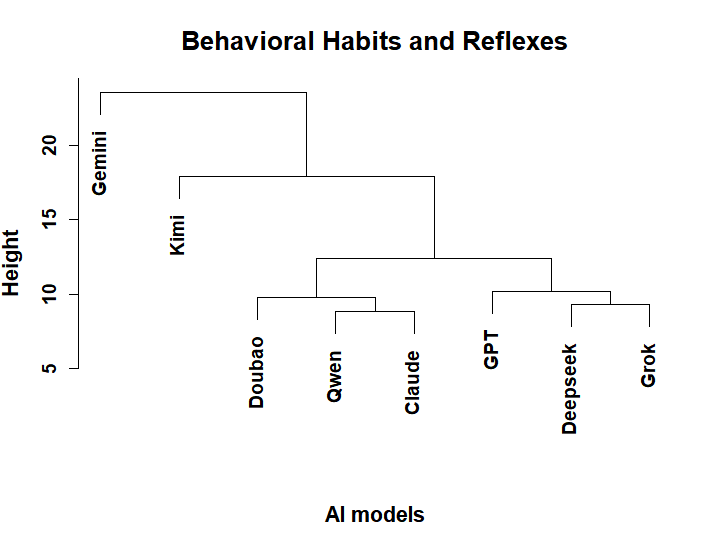

**Figure S16. The clustering analysis of Behavioral habits and reflexes scores in 8 AI models.**

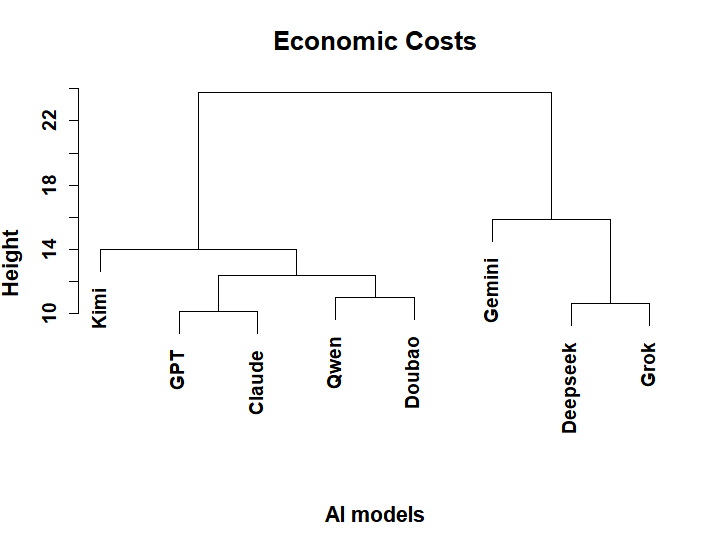

**Figure S17. The clustering analysis of Economic costs scores in 8 AI models.**
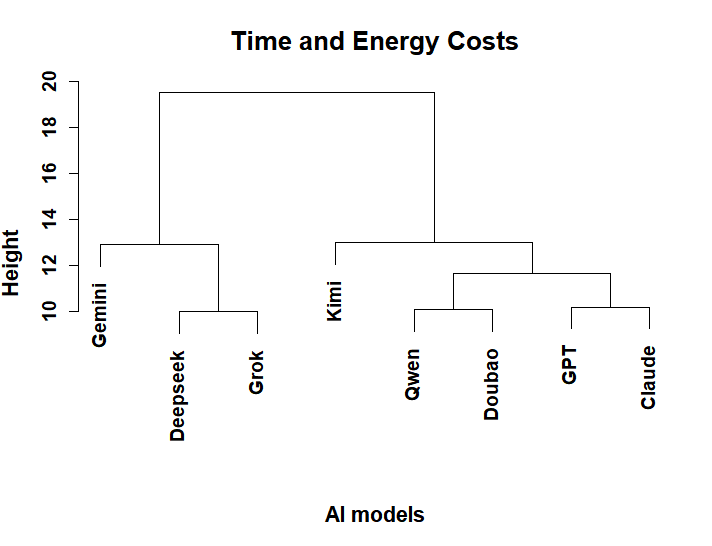

**Figure S18. The clustering analysis of Time and energy costs scores in 8 AI models.**

5. **The unequal difficulty of high BMI intervention in 18 indicators.**

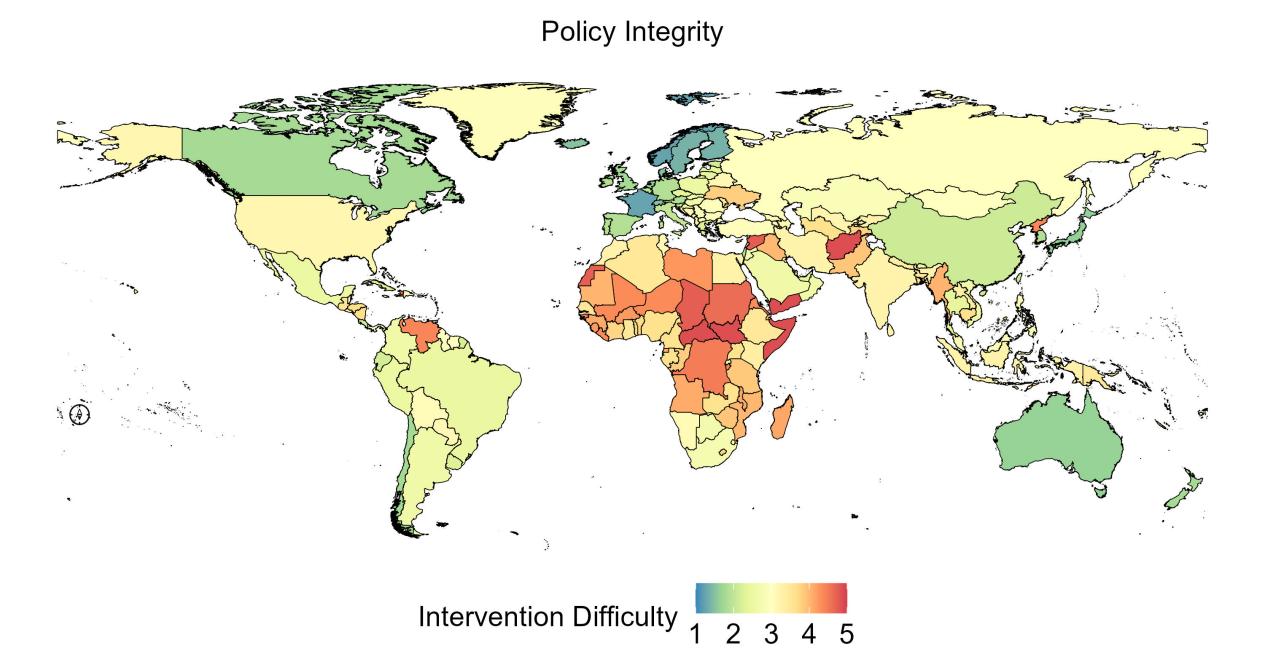

**Figure S19. Global unequal difficulty of high BMI intervention in Policy integrity.**

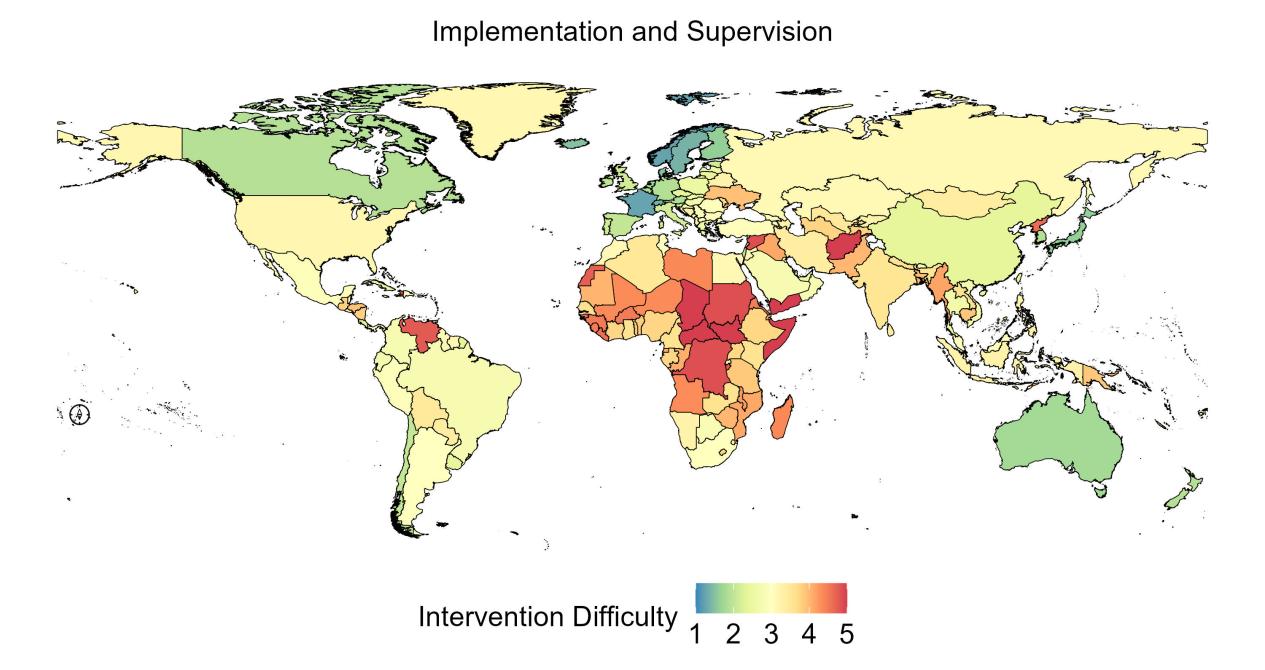

**Figure S20. Global unequal difficulty of high BMI intervention in Implementation and supervision.**

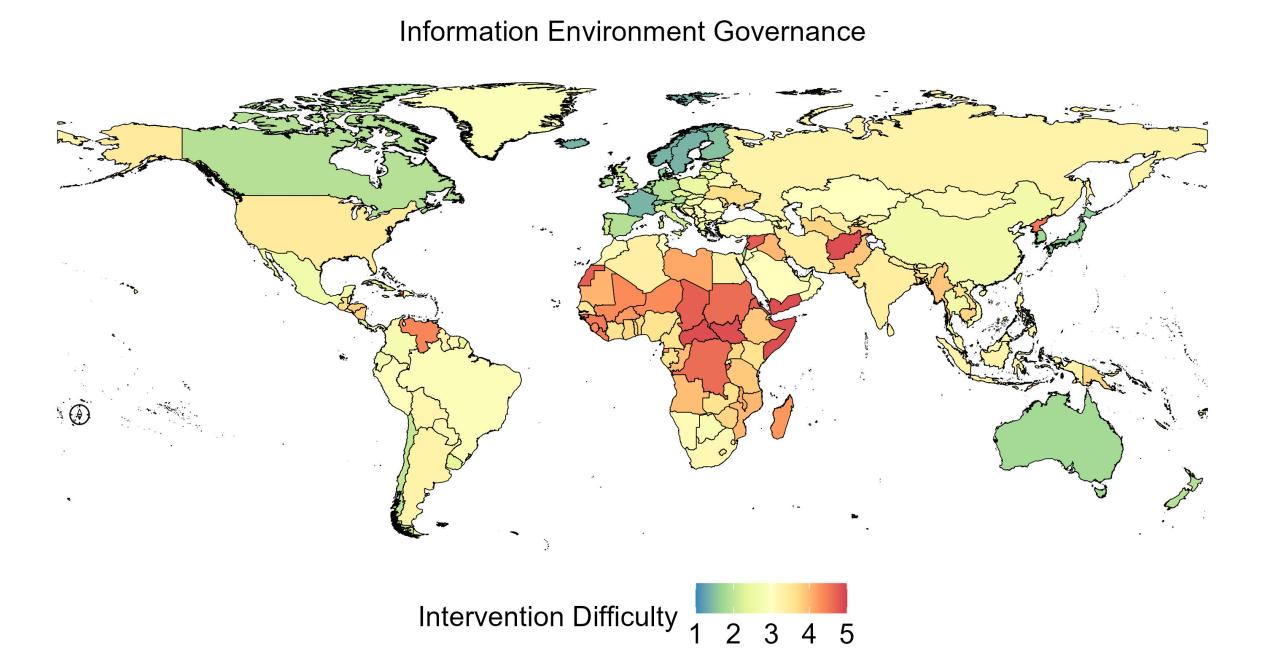

**Figure S21. Global unequal difficulty of high BMI intervention in Information environment governance.**

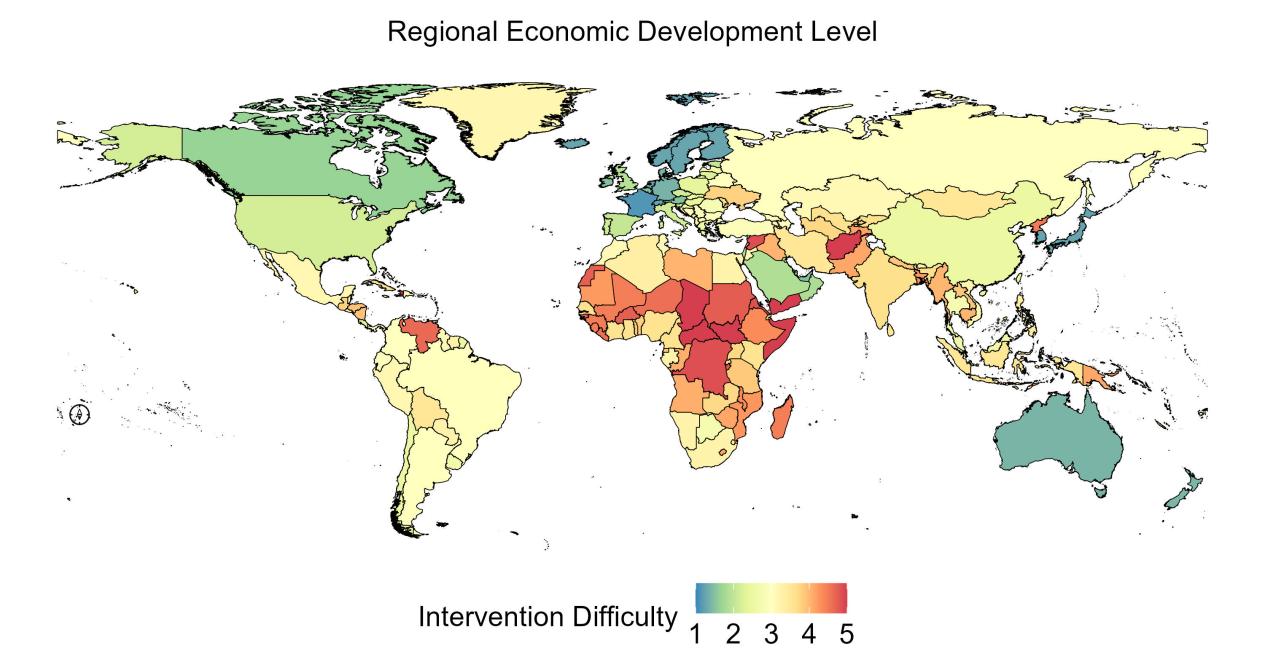

**Figure S22. Global unequal difficulty of high BMI intervention in Regional economic development level.**

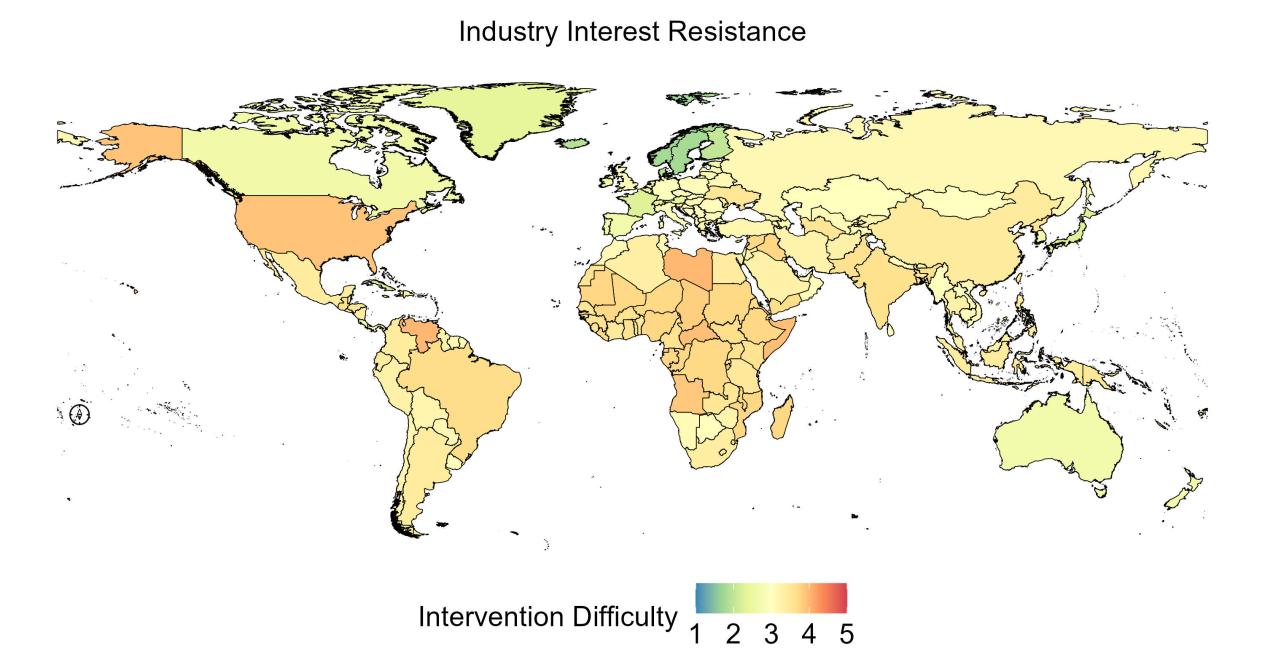

**Figure S23. Global unequal difficulty of high BMI intervention in Industry interest resistance.**

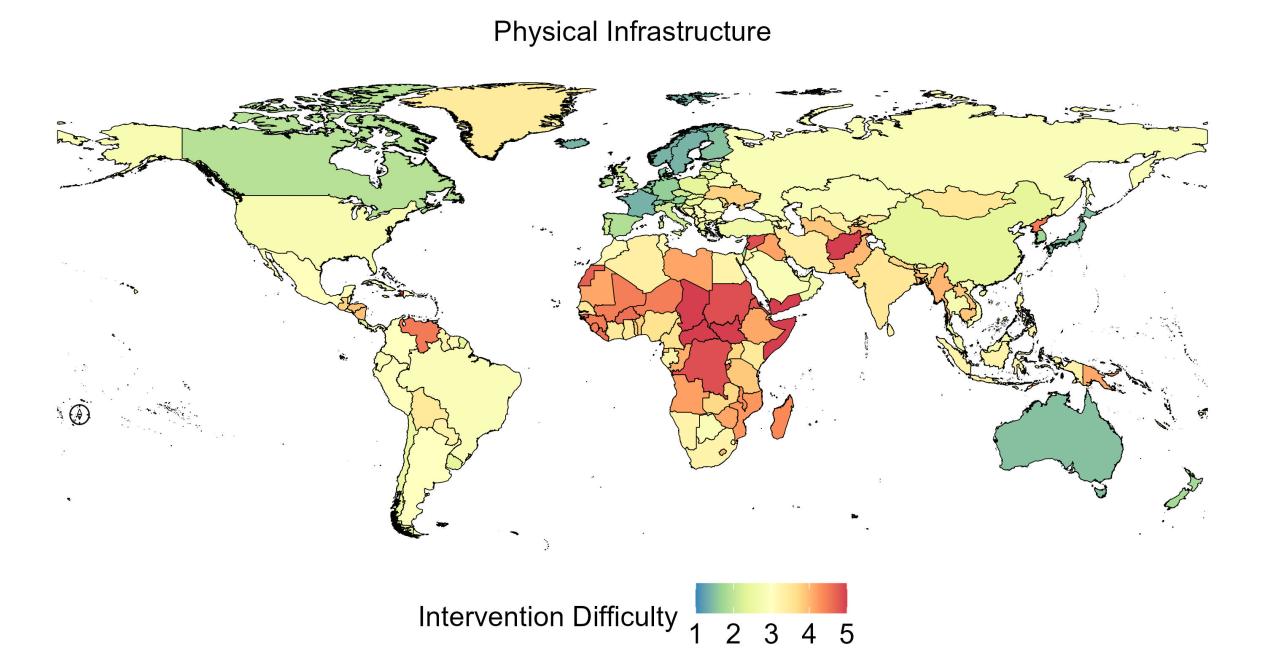

**Figure S24. Global unequal difficulty of high BMI intervention in Physical infrastructure.**

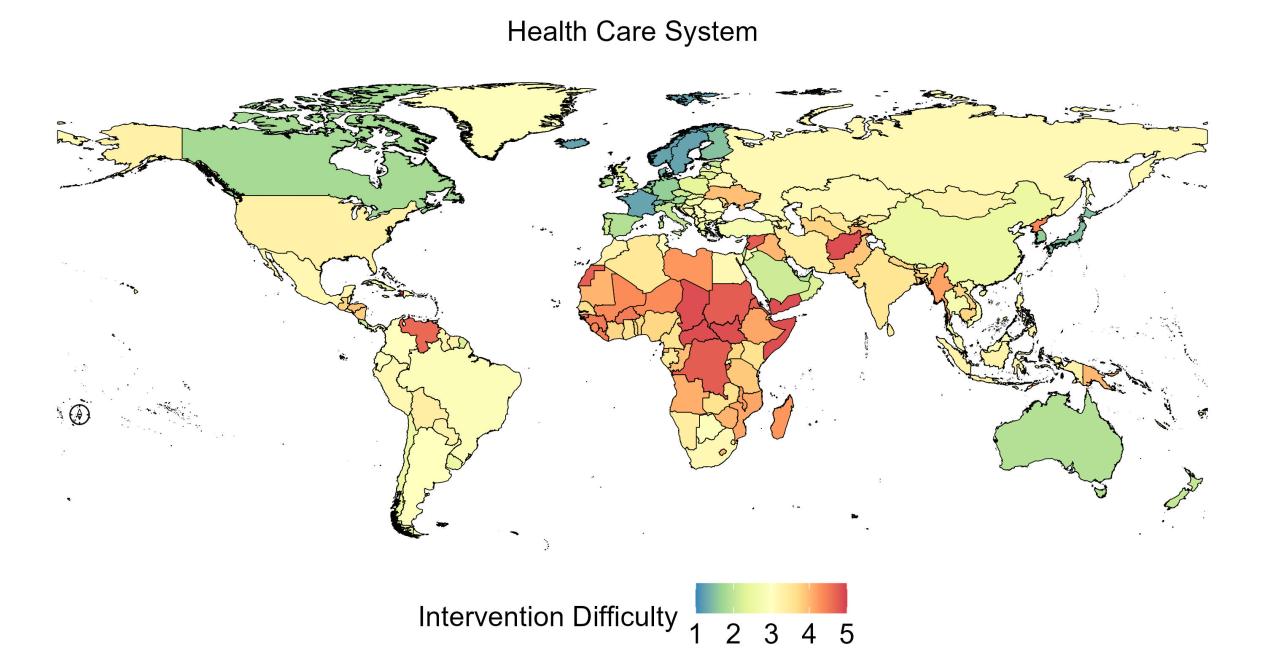

**Figure S25. Global unequal difficulty of high BMI intervention in Health care system**

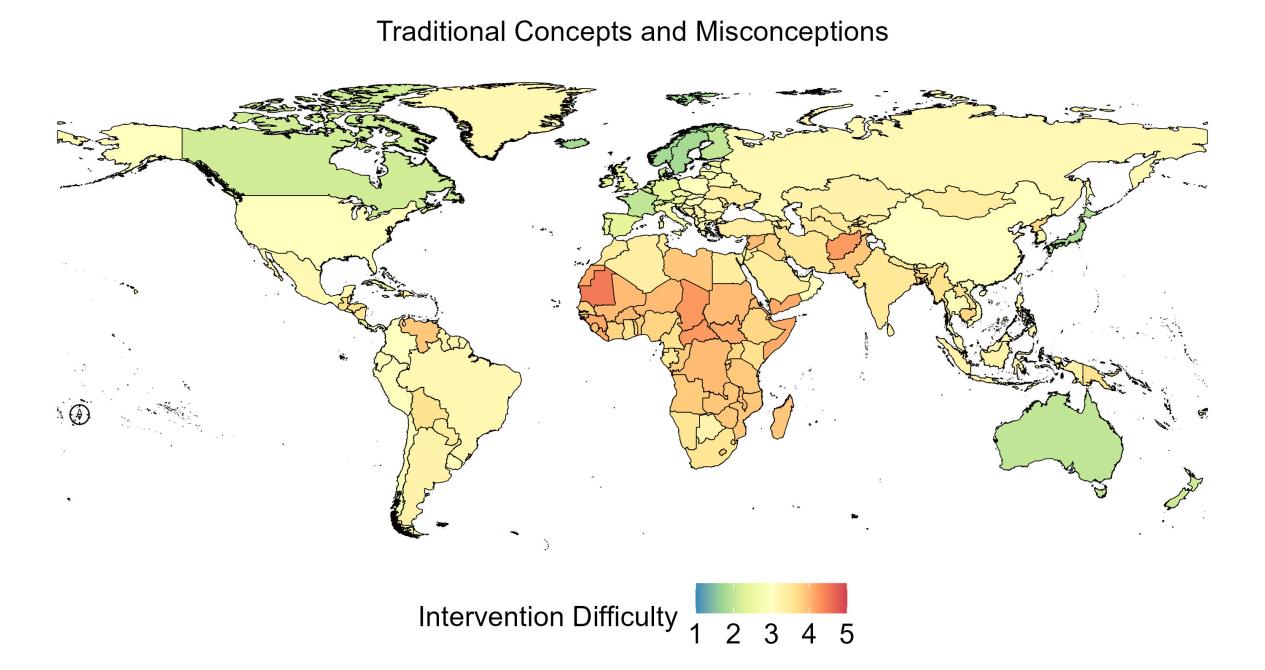

**Figure S26. Global unequal difficulty of high BMI intervention in Traditional concepts and misconceptions.**

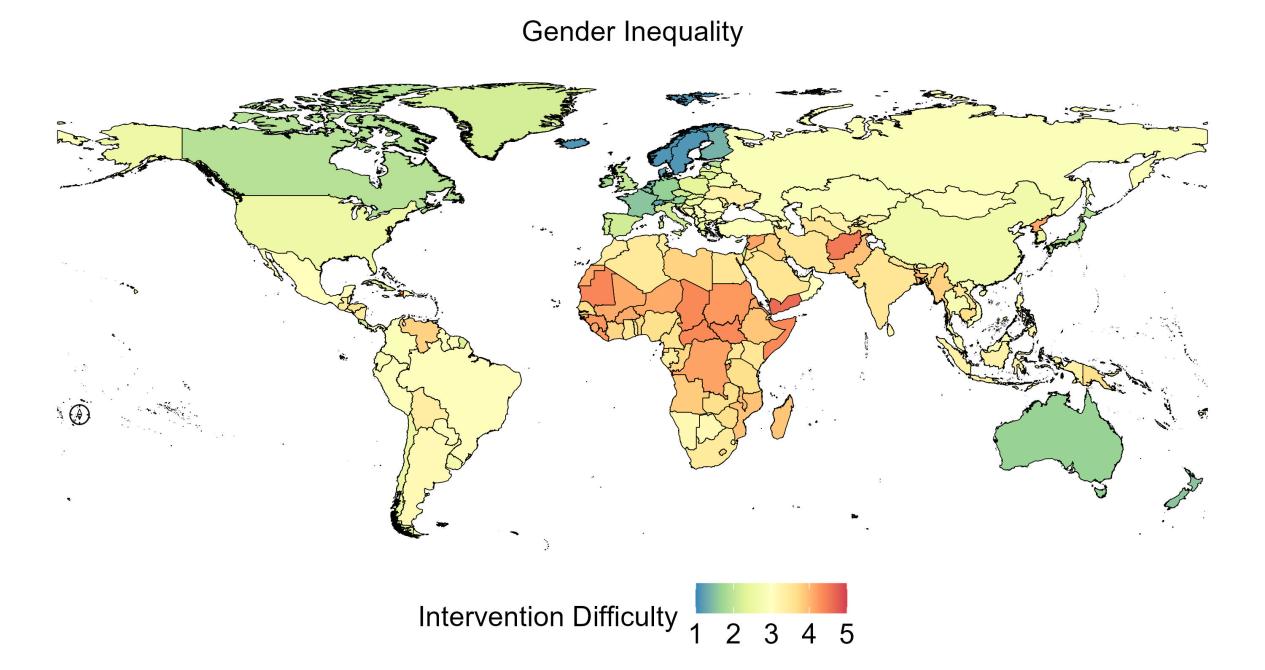

**Figure S27. Global unequal difficulty of high BMI intervention in Gender inequality.**

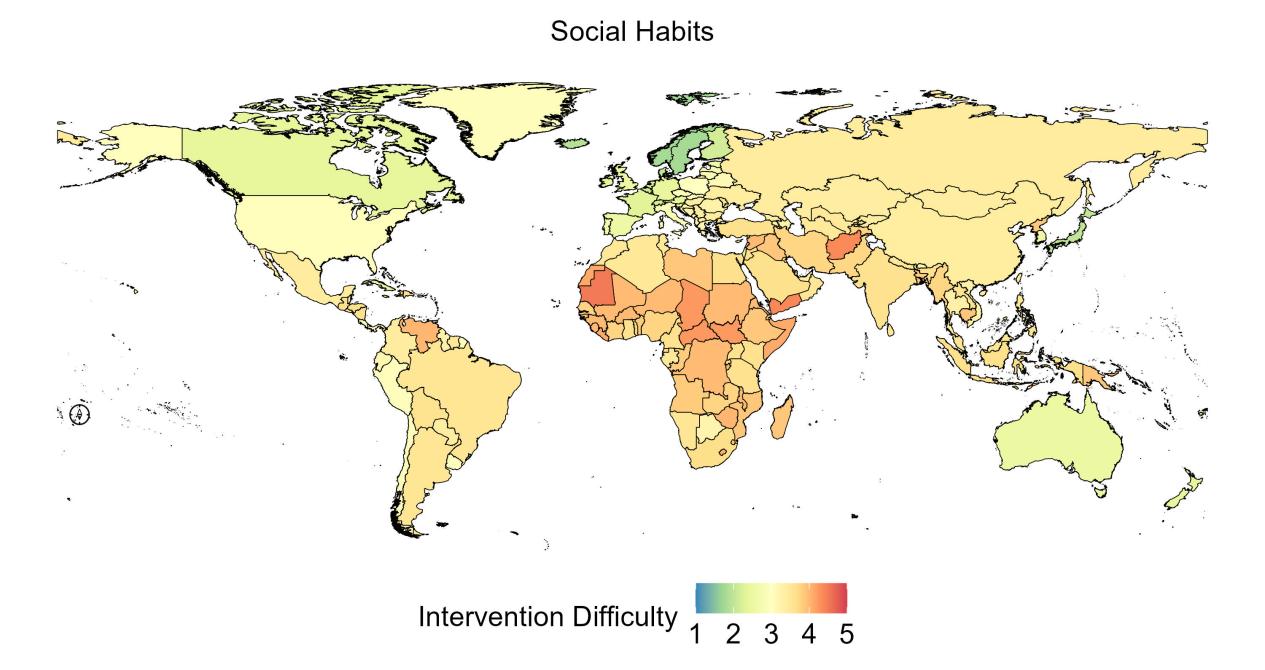

**Figure S28. Global unequal difficulty of high BMI intervention in Social habits.**

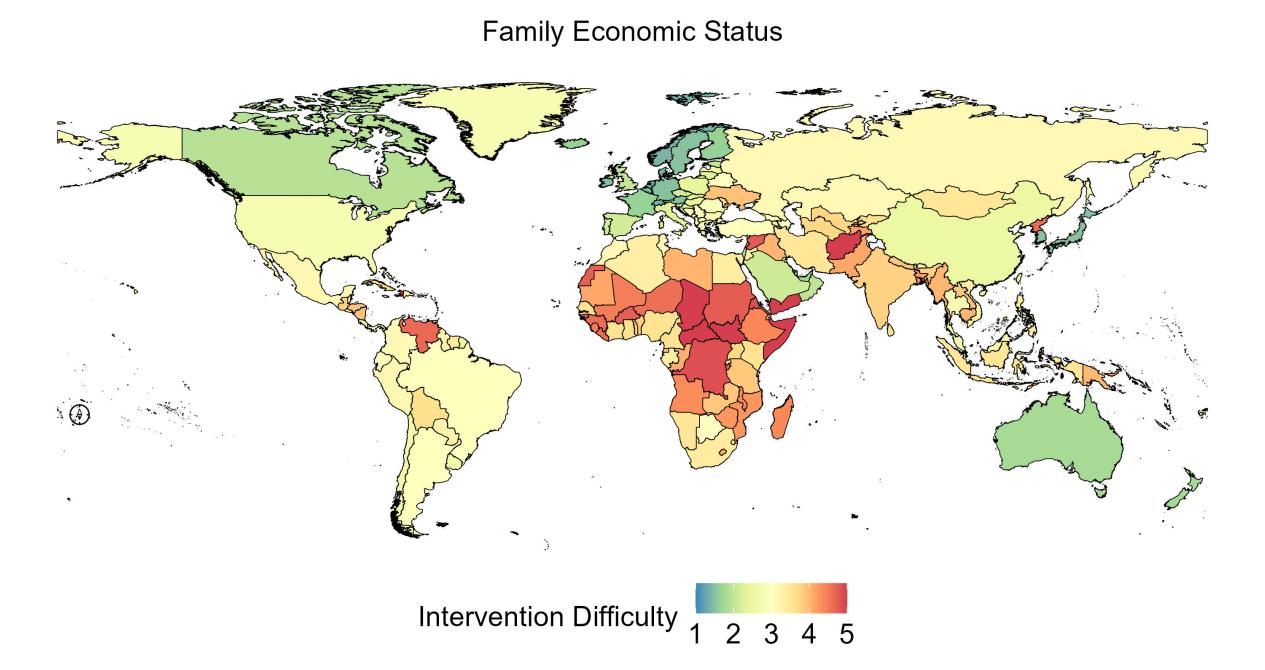

**Figure S29. Global unequal difficulty of high BMI intervention in Family economic status.**

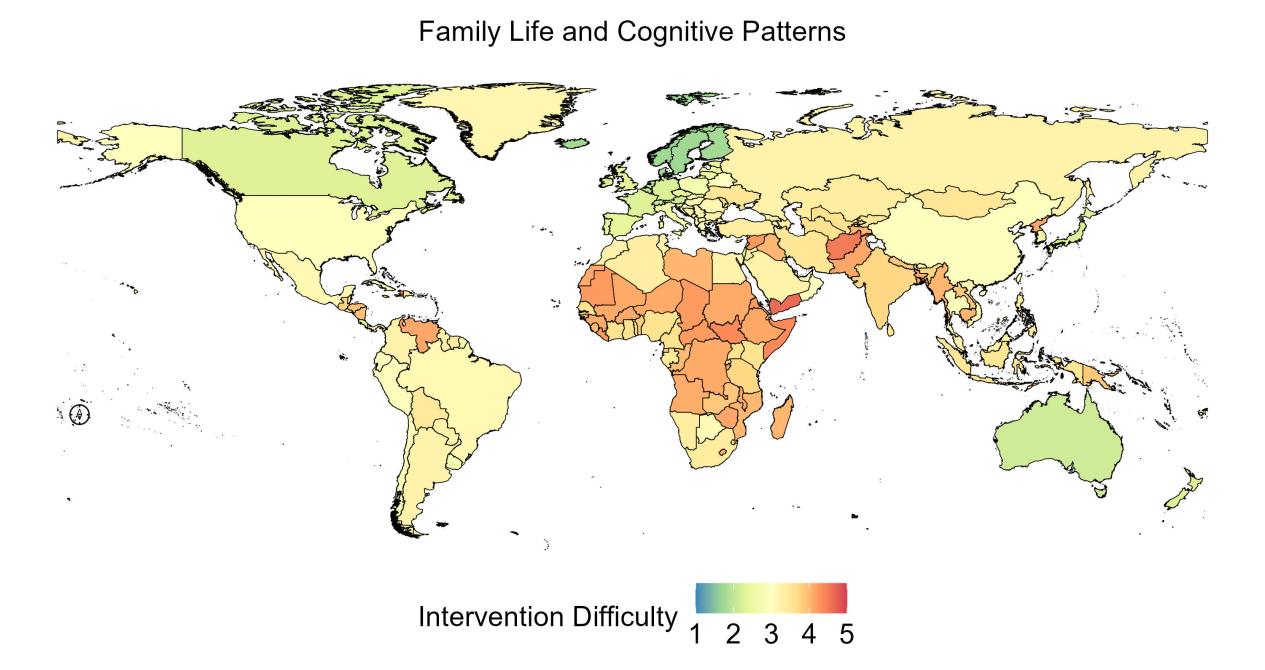

**Figure S30. Global unequal difficulty of high BMI intervention in Family life and cognitive patterns.**

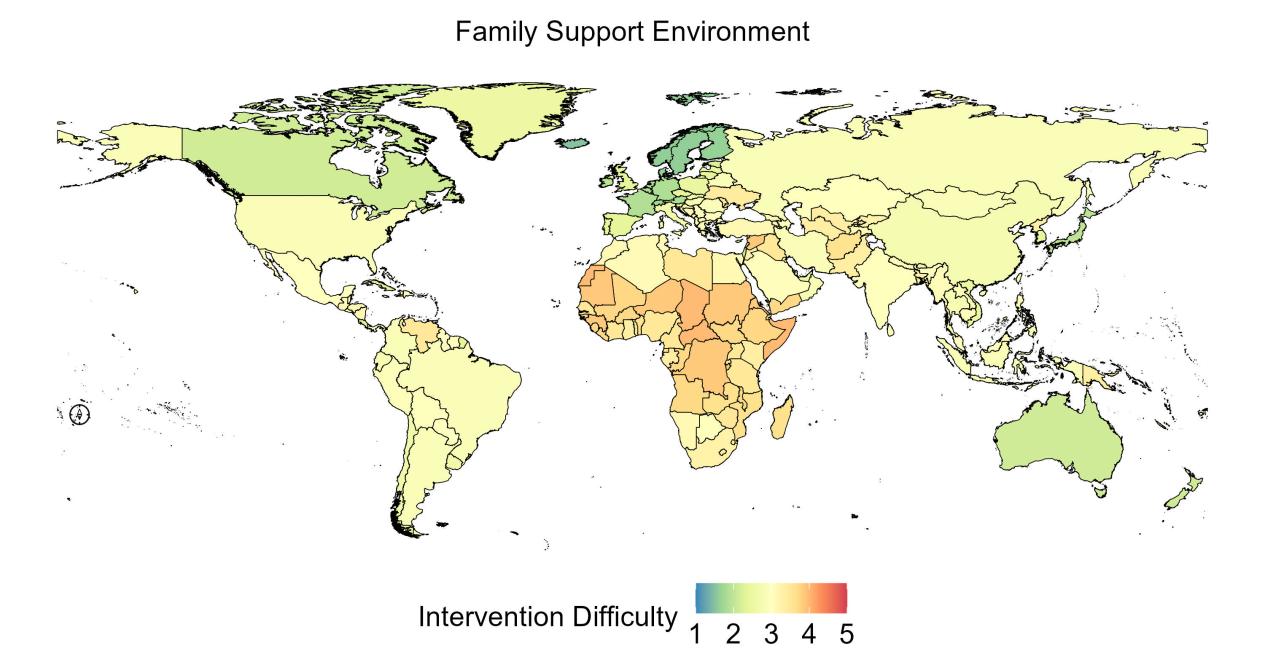

**Figure S31. Global unequal difficulty of high BMI intervention in Family support environment.**

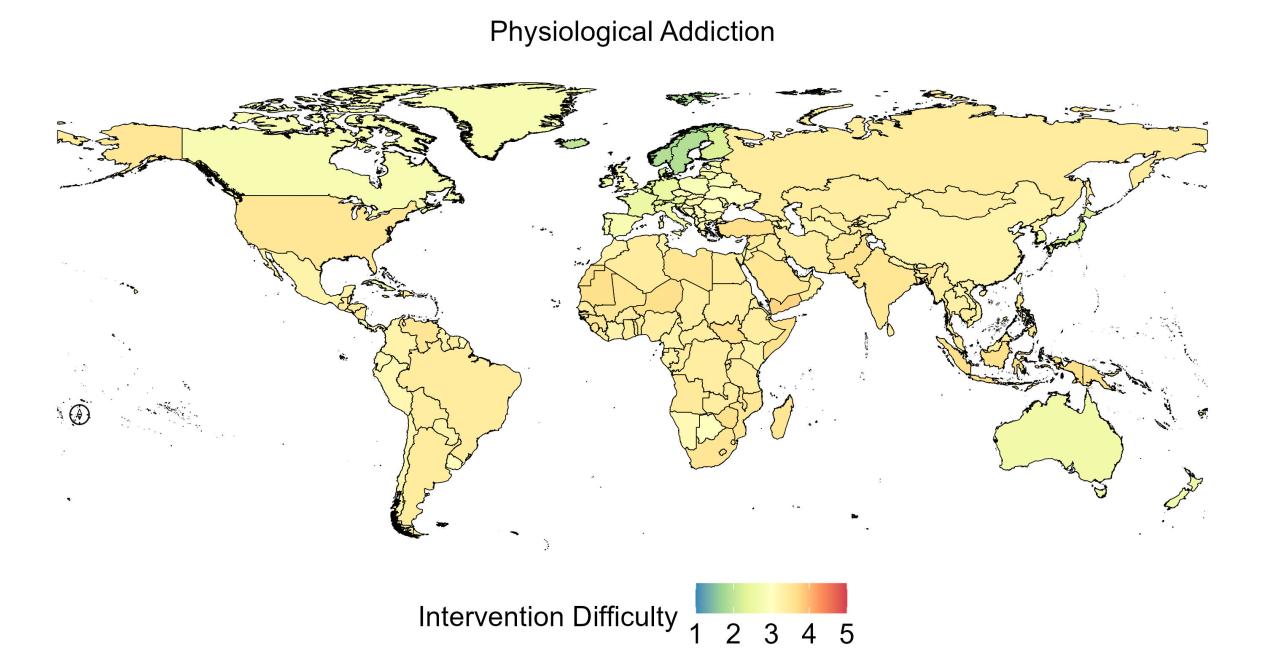

**Figure S32. Global unequal difficulty of high BMI intervention in Physiological addiction.**

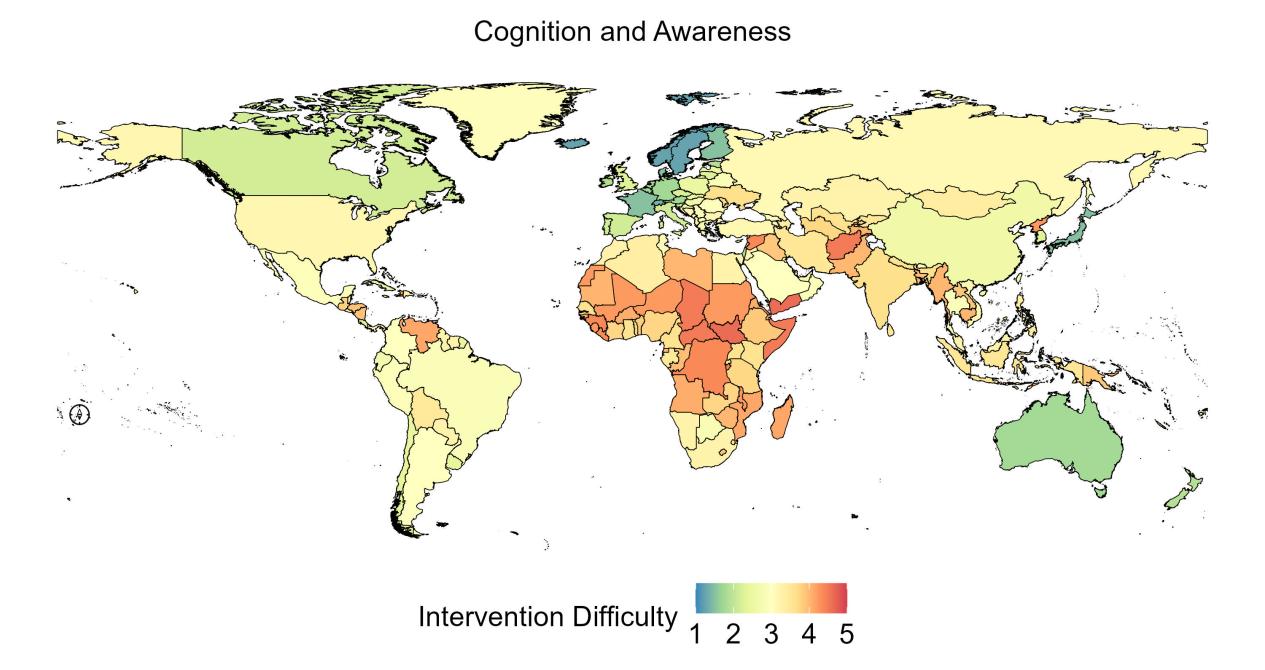

**Figure S33. Global unequal difficulty of high BMI intervention in Cognition and awareness.**

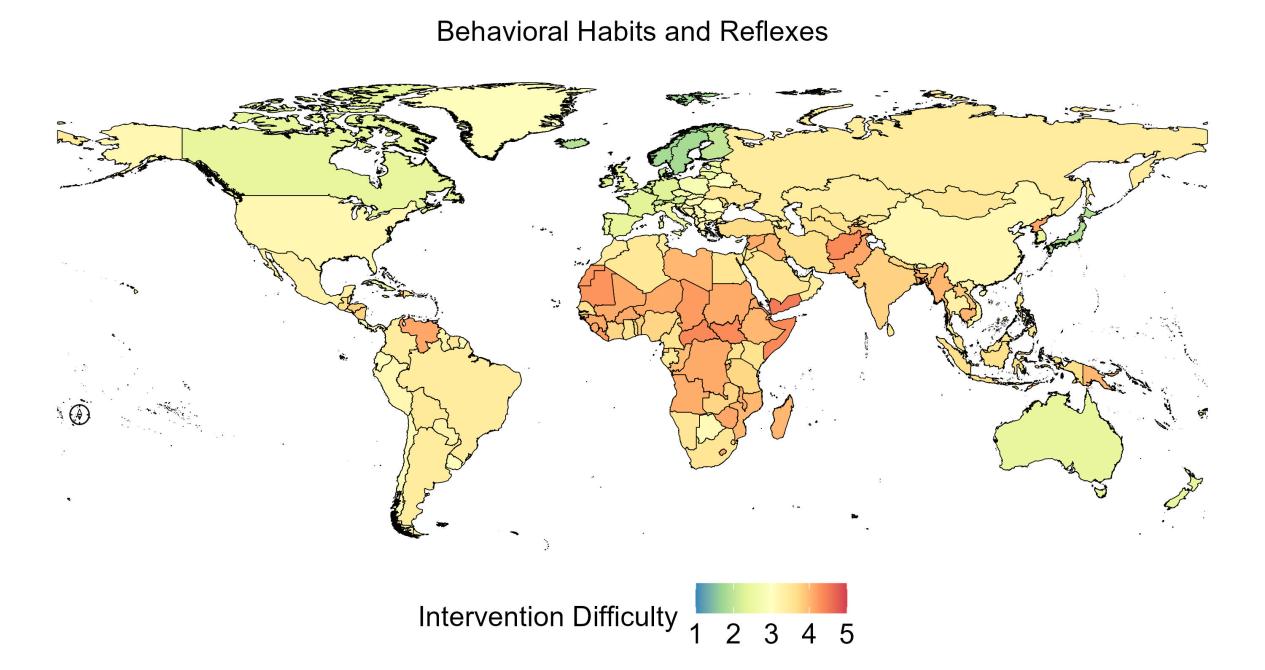

**Figure S34. Global unequal difficulty of high BMI intervention in Behavioral habits and reflexes.**

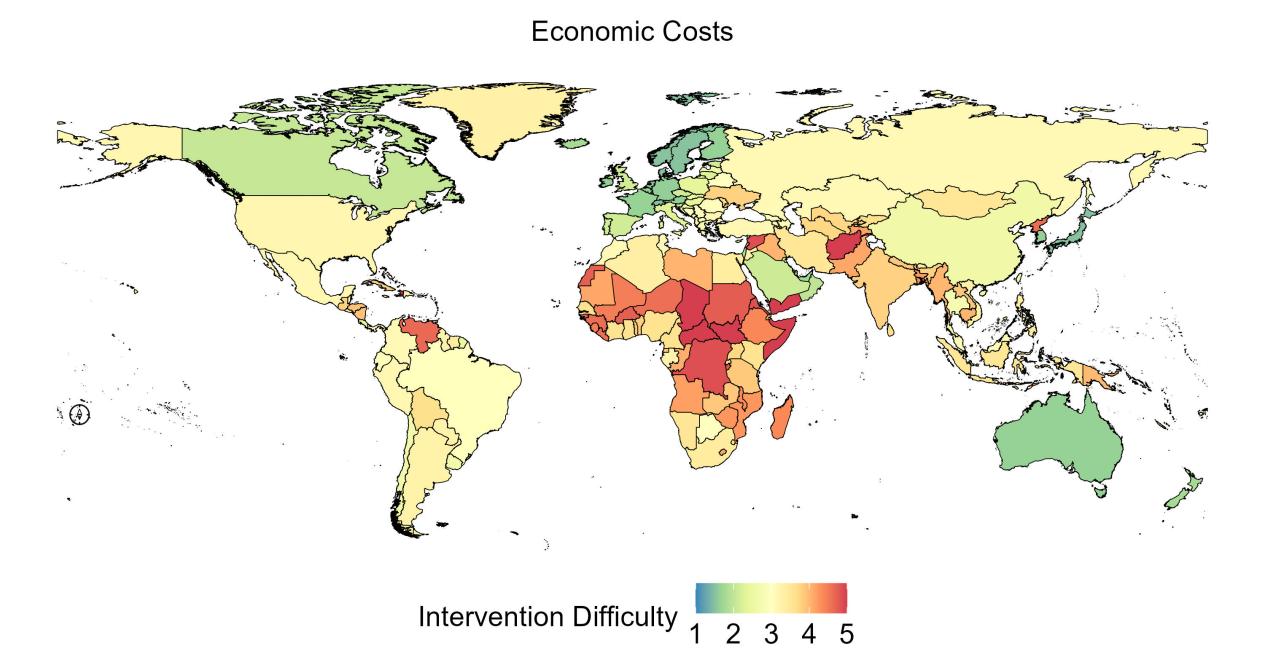

**Figure S35. Global unequal difficulty of high BMI intervention in Economic costs.**

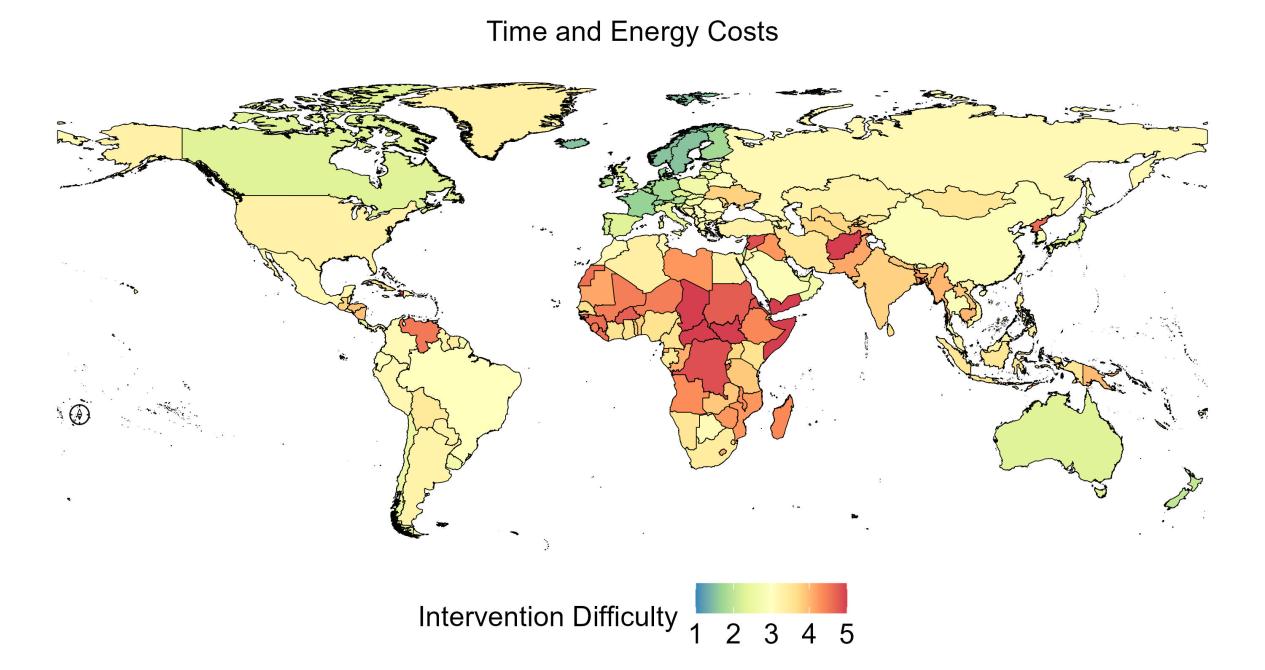

**Figure S36. Global unequal difficulty of high BMI intervention in Time and energy costs.**

6. **Interactive website provides user-friendly access for global difficulty of high BMI intervention.**

To enable a thorough investigation of the global difficulty of high BMI intervention, we created an interactive website (http://www.deepburden.com/high-bmi) that offers user-friendly access to comprehensive results. The website comprises four modules cataloging: *Home*, *GAME framework*, *Result*, and *Contact*. We provided the introduction of GAME framework at the *GAME framework* page.

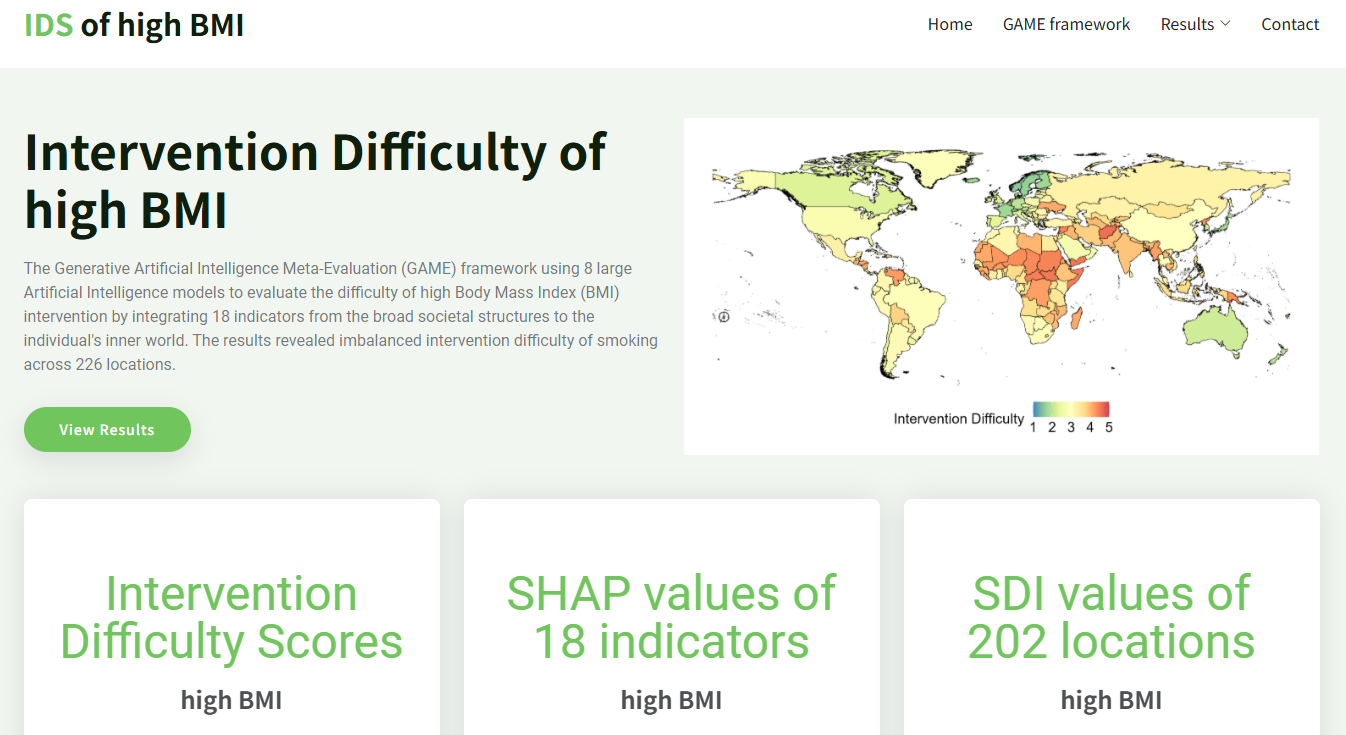

**Figure S37. Homepage of the website.**

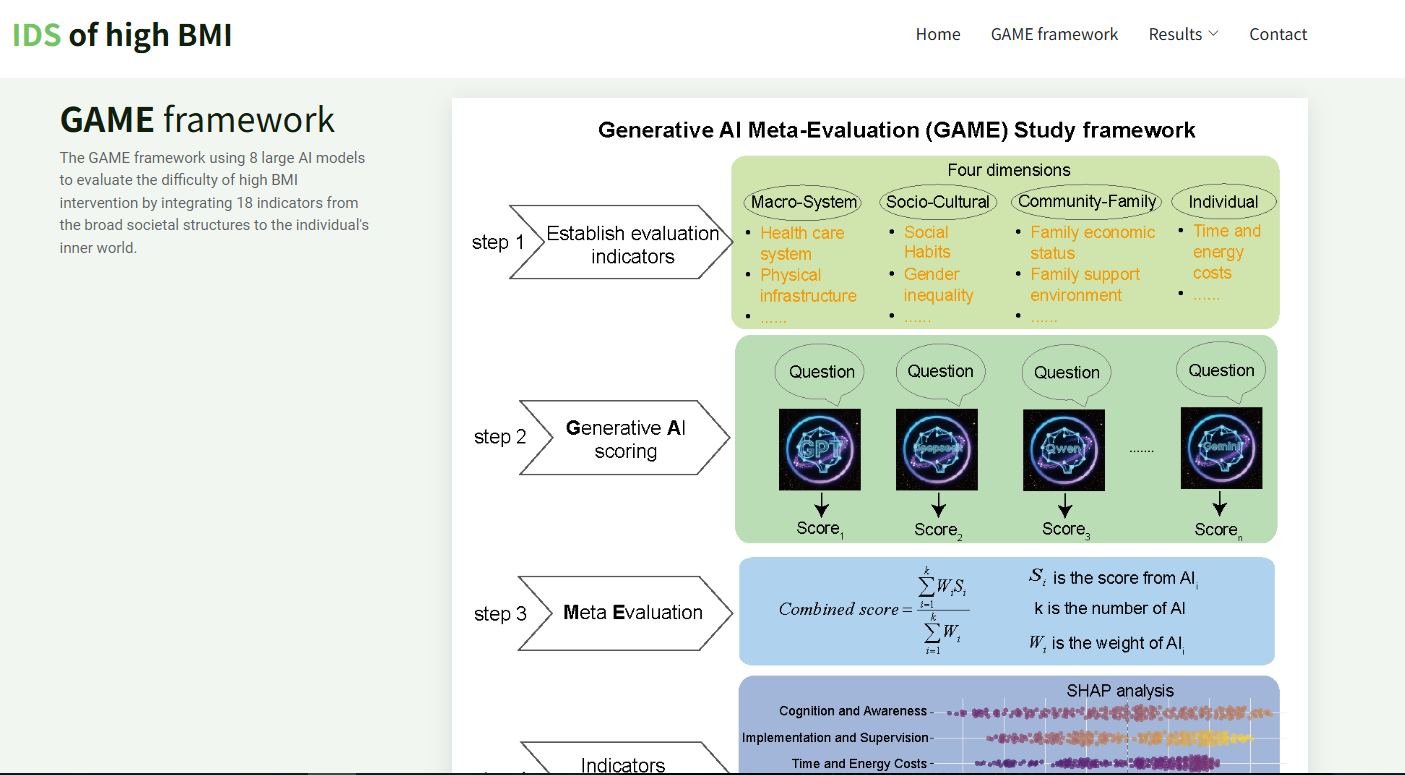

**Figure S38. GAME framework of the website.**

There were three parts in Result page, including “*Intervention Difficulty Scores*”, “*SHAP values of 18 indicators*”, and “*SDI values of 202 locations*”. The intervention difficulty scores of high BMI in 226 locations were in *Intervention Difficulty Scores* page (Figure S39). The SHapley Additive exPlanation (SHAP) values indicating the impact of 18 indicators were provided in *SHAP values of 18 indicators* page (Figure S40). In addition, we offered the SDI values and levels for 202 locations obtained from Global Burden of Diseases 2021 study in *SDI values of 202 locations* page (Figure S41).

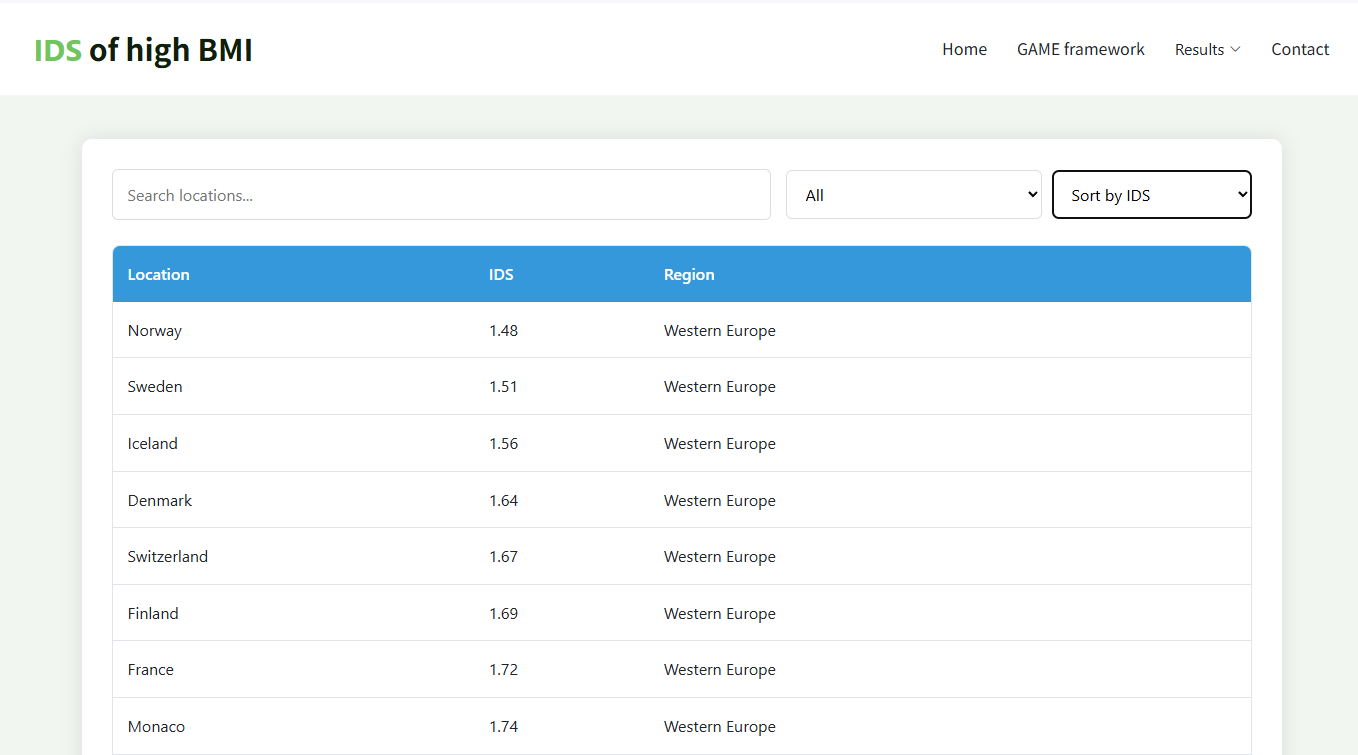

**Figure S39. Results of “Intervention Difficulty Scores” page.**

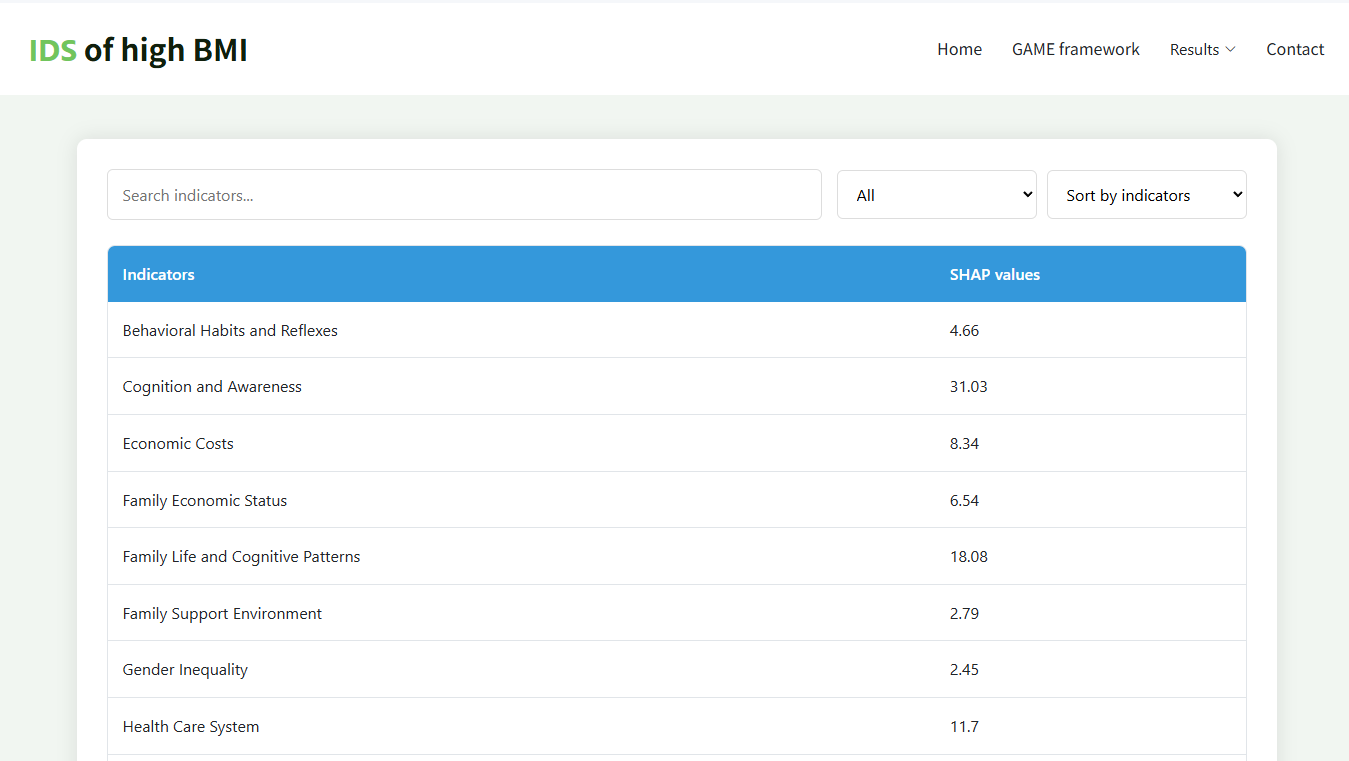

**Figure S40. Results of “SHAP values of 18 indicators” page.**

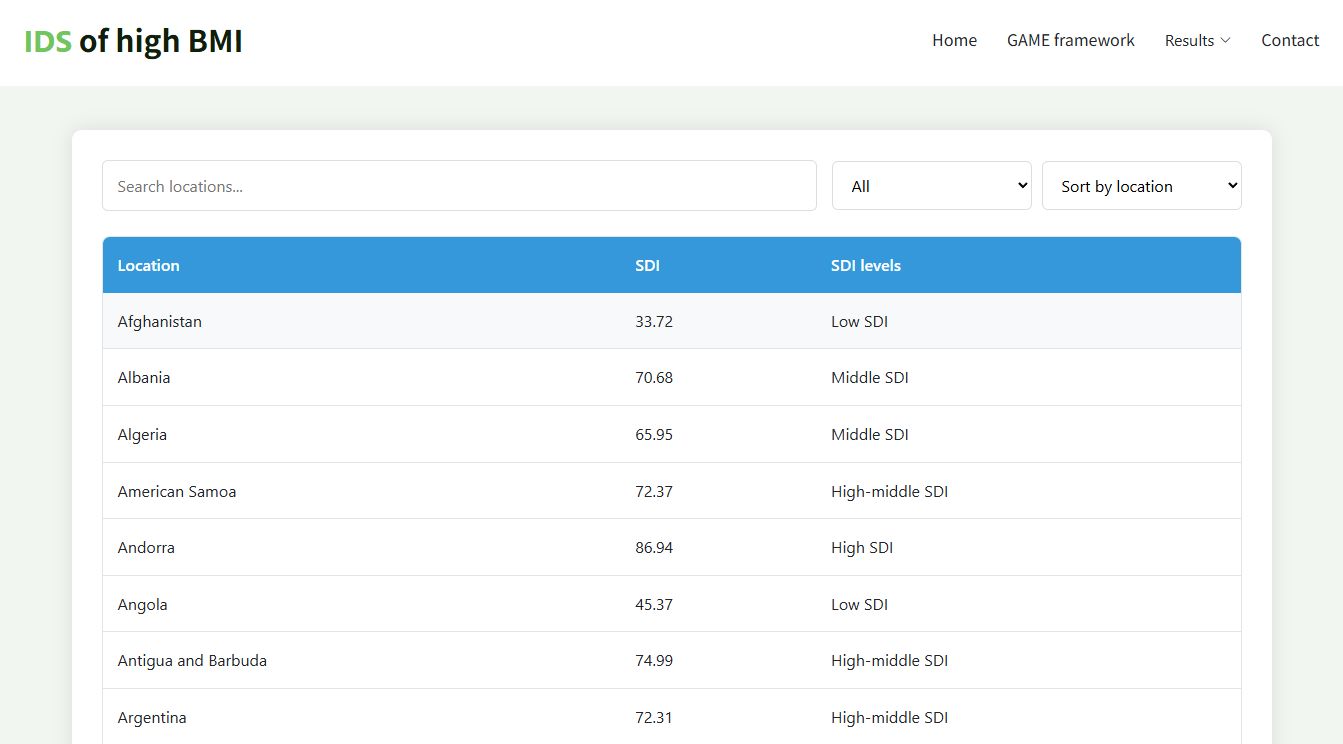

**Figure S41. Results of “SDI values of 202 locations” page.**
